# Androgens mediate sexual dimorphism in Pilarowski-Bjornsson Syndrome

**DOI:** 10.1101/2025.05.06.25326635

**Authors:** Kimberley Jade Anderson, Eirny Tholl Thorolfsdottir, Ilana M. Nodelman, Sara Tholl Halldorsdottir, Stefania Benonisdottir, Malak Alghamdi, Naif Almontashiri, Brenda J. Barry, Matthias Begemann, Jacquelyn F. Britton, Sarah Burke, Benjamin Cogne, Ana S.A. Cohen, Carles de Diego Boguñá, Evan E. Eichler, Elizabeth C. Engle, Jill A. Fahrner, Laurence Faivre, Mélanie Fradin, Nico Fuhrmann, Christine W. Gao, Gunjan Garg, Dagmar Grečmalová, Mina Grippa, Jacqueline R. Harris, Kendra Hoekzema, Tova Hershkovitz, Sydney Hubbard, Katrien Janssens, Julie A. Jurgens, Stanislav Kmoch, Cordula Knopp, Meral Aktas Koptagel, Farah A. Ladha, Pablo Lapunzina, Tobias Lindau, Marije Meuwissen, Andreina Minicucci, Emily Neuhaus, Mathilde Nizon, Lenka Nosková, Kristen Park, Chirag Patel, Rolph Pfundt, Pankaj Prasun, Nils Rahner, Nathaniel H. Robin, Carey Ronspies, Jasmin Roohi, Jill Rosenfeld, Margarita Saenz, Carol Saunders, Zornitza Stark, Isabelle Thiffault, Sarah Thull, Danita Velasco, Clara Velmans, Jolijn Verseput, Antonio Vitobello, Tianyun Wang, Karin Weiss, Ingrid M. Wentzensen, Genay Pilarowski, Thor Eysteinsson, Madelyn Gillentine, Kári Stefánsson, Agnar Helgason, Gregory D. Bowman, Hans Tomas Bjornsson

## Abstract

Sex-specific penetrance in autosomal dominant Mendelian conditions is largely understudied. The neurodevelopmental disorder Pilarowski-Bjornsson syndrome (PILBOS) was initially described in females. Here, we describe the clinical and genetic characteristics of the largest PILBOS cohort to date, showing that both sexes can exhibit PILBOS features, although males are overrepresented. A mouse model carrying a human-derived *Chd1* missense variant (*Chd1^R^*^616^*^Q^*^/+^) displays female-restricted phenotypes, including growth deficiency, anxiety and hypotonia. Orchiectomy unmasks a growth deficiency phenotype in male *Chd1^R^*^616^*^Q^*^/+^ mice, while testosterone rescues the phenotype in females, implicating androgens in phenotype modulation. In the gnomAD and UK Biobank databases, rare missense variants in *CHD1* are overrepresented in males, supporting a male protective effect. We identify 33 additional highly constrained autosomal genes with missense variant overrepresentation in males. Our results support androgen-regulated sexual dimorphism in PILBOS and open novel avenues to understand the mechanistic basis of sexual dimorphism in other autosomal Mendelian disorders.

**Graphical Abstract:** 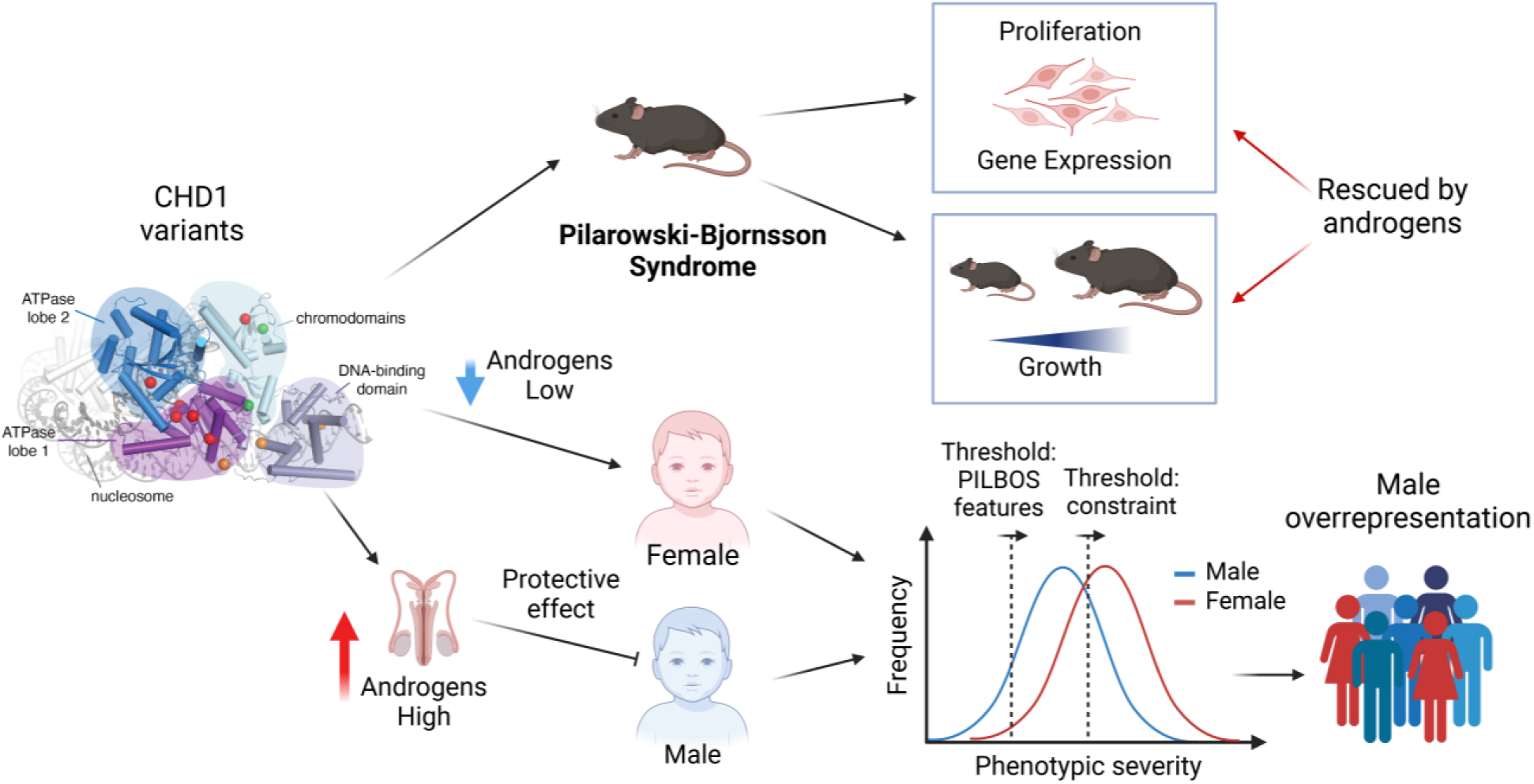

## Introduction

Traditionally, autosomal dominant disorders have been considered to have limited sex differences, unlike X-linked disorders defined by this feature. Pilarowski-Björnsson syndrome (PILBOS, MIM#617682) is an autosomal dominant disorder first described by our group in 2018 in six affected girls with developmental delay, hypotonia, speech apraxia, autism, and seizures^1^. Each individual carried a heterozygous missense variant in the chromatin remodeler gene *CHD1*, located on chromosome 5, and four were de novo variants. The absence of males led to the hypothesis of male lethality, consistent with the strong male overrepresentation in neurodevelopmental disorders (NDDs)^2,3^. Since then, two case reports have been published including individuals of either sex^4,5^. Thus, the question arises: does this disorder affect both sexes equally and does the disease mechanism of these variants involve gain-of-function or loss-of-function? To date no large scale PILBOS study has been performed and therefore the full phenotypic details and mechanistic basis remains elusive.

CHD1 is a highly conserved chromatin remodeler that was first characterized for its role at sites of active transcription^6,7^, performing ATP-dependent chromatin remodeling of nucleosomes^8,9^. CHD1 is known to recognize and directly bind trimethylated histone H3 lysine 4 (H3K4me3)^10^ and is thought to drive global transcriptional output^11^, co-operating with elongation factors^12^ and shifting the +1 nucleosome to allow RNA polymerase II to escape from promoters^13^. CHD1 also has important roles in DNA replication, negatively regulating S-phase in yeast^14^, remodeling the nucleosome behind the replication fork^15^, and remodeling chromatin at stalled replication forks^16^. Furthermore, CHD1 is important for the homologous recombination and nucleotide excision repair DNA damage repair pathways and is known to directly interact with DNA repair factors^17–21^.

The investigation of sex differences in mouse models of neurological conditions is critically understudied^22^. The careful identification of robust examples of sexually dimorphic conditions could help identify additional such disorders and lead to therapeutic strategies for those sharing the same causal modifier. An obvious difference between the two sexes involves the sex hormones, which play key roles in neurodevelopment as the main drivers of sex differences in the brain^23^. Interestingly, in prostate cancer, CHD1 is known to directly interact with the androgen receptor (AR), binding to AR-occupied enhancers, and loss of CHD1 leads to genome-wide reorganization of AR binding^24,25^. The CHD1-AR interaction may offer a unique window into a potential mechanism for sexual dimorphism in PILBOS and other related NDD phenotypes.

Here, we provide a comprehensive characterization of a large set of individuals with a diagnosis of PILBOS and their corresponding *CHD1* genetic variants. Using a novel mouse model (*Chd1^R^*^616^*^Q/+^*) carrying a variant identified in one of these individuals, we demonstrate sexual dimorphism of core PILBOS phenotypes. We further show that exposure to androgens mediates the protective effect observed in males. Finally, we utilize human population data to demonstrate differential frequencies of *CHD1* variants between the sexes and show that our findings of sexual dimorphism may extend to other autosomal dominant Mendelian disorders.

## Results

### PILBOS affects both sexes through distinct mutational mechanisms

To further explore the phenotypic presentation of PILBOS, we identified additional individuals using GeneMatcher^26^ (https://genematcher.org/), through direct communication with individuals, families and providers, and through an online PILBOS support group. This resulted in the identification of 48 unrelated individuals from 14 countries. Together with the eight individuals previously reported in the literature^1,4,5,27–29^, this amounted to 52 separate variants in 56 individuals: 39 males and 17 females.

Notably, males were significantly overrepresented in this cohort (*p* = 0.0046, binomial test). A total of 33 individuals carried 29 missense variants, of which: 17 were *de novo* (52%), 5 inherited (15%), and 11 unknown (33%). Twenty individuals carried truncating loss of function (LOF) variants: 11 were *de novo* (55%), 4 inherited (20%), and 5 unknown (25%); one carried a stop-loss variant (*de novo*), and two carried possible non-canonical splice-site variants (one *de novo*, one of unknown inheritance). Three of the missense variants were recurring: firstly, a variant identified in two unrelated males (c.2090G>A, p.Arg697Gln), secondly, a variant identified in two unrelated females (c.3335G>T, p.Arg1112Leu), and thirdly, a variant identified in two sisters and an unrelated male (c.5123G>A, p.Arg1708Gln). This third variant (c.5123G>A, p.Arg1708Gln) has been reported at a low frequency (3.73 × 10^-6^) in the gnomAD population database (v4.1.0)^30^ and has also been identified in unaffected parents from clinical cohorts, raising the possibility of this being a benign variant. However, since we are interested in penetrance, here we include information on all variants.

Of all the variants, 24 (46%) were classified as being likely pathogenic using the American College of Medical Genetics and Genomics (ACMG) guidelines. Most of the missense variants (83%) were classified as variants of unknown significance, with five of them (17%) being classified as likely pathogenic. We used AlphaMissense^31^ to perform structural predictions, identifying 12 additional missense variants that may disrupt structure and function (deleterious or uncertain classification, see **Supplementary Table 1**), and created a color-coded visual representation of LOF and missense variants for either sex (**Figure 1A**). Twelve of the 29 missense variants (41%) are categorized as deleterious and appear to lead to loss of protein function based on AlphaMissense structural predictions, and a further four missense variants (14%) are categorized as uncertain, while 13 are categorized as benign (45%). Comparing the sexes, 21 missense variants were found in males (64%) compared to 12 in females (36%). For the 21 individuals carrying putatively disruptive missense variants (categorized as deleterious or uncertain), there were 11 males and 10 females, whereas 15 males carried LOF truncating variants categorized as likely pathogenic (79%) compared to 4 females (21%) (**Fig. 1B**). Subsequently, our collaborators at GeneDx analyzed a separate cohort of individuals from their database who underwent exome sequencing-based testing and were found to have pathogenic or likely pathogenic *CHD1* variants (n < 15). Notably, no females in this cohort had frameshift or nonsense variants. Instead, females had missense (67%) or splice site variants (33%), while males had LOF (33%) or missense variants (67%).

**Figure 1:**
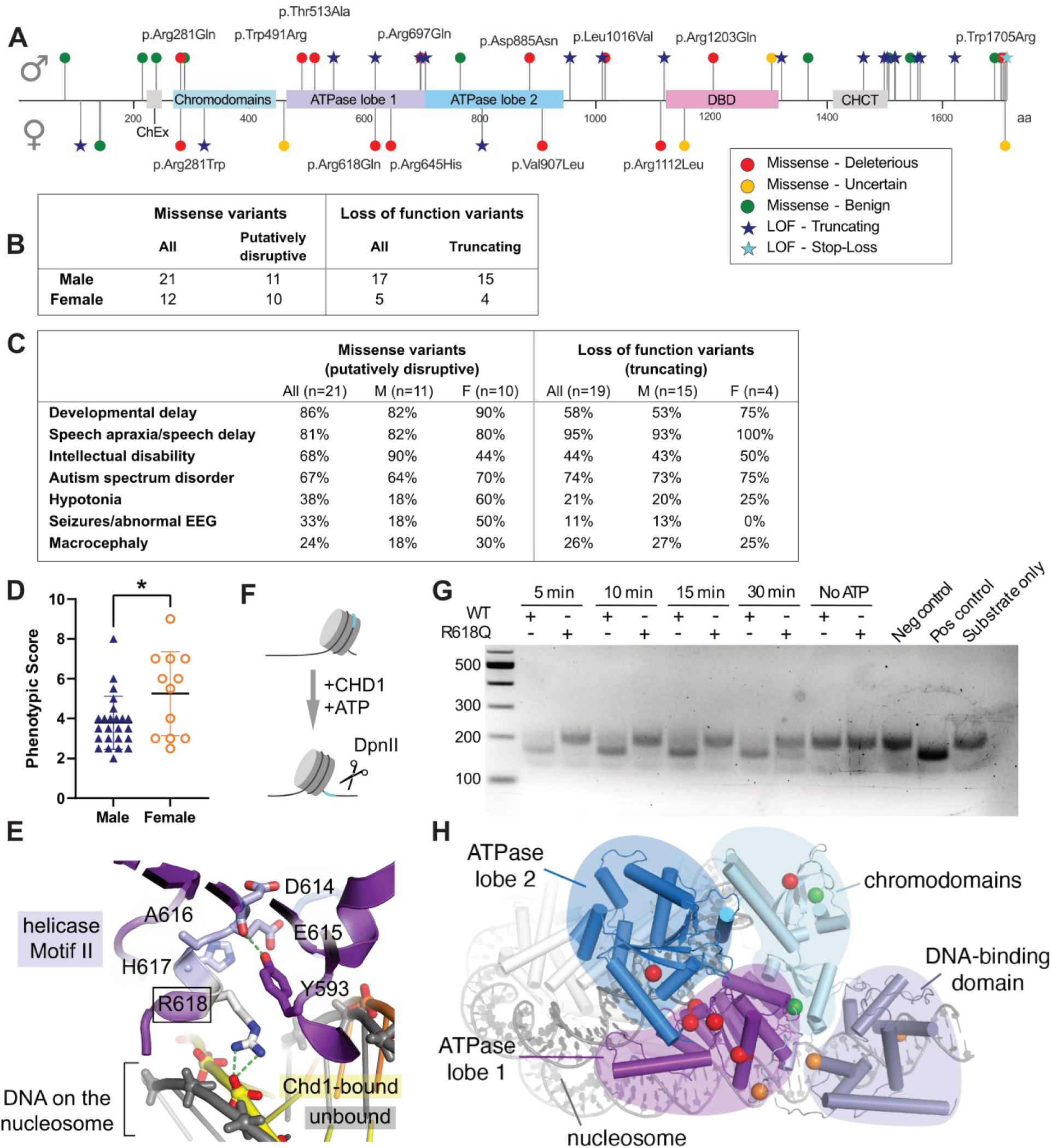
PILBOS missense variants lead to loss of function of *CHD1*. (A) A schematic representation of the CHD1 protein. Location of variants shown as stars (LOF variants) and circles (missense variants). Male variants are above and female variants below the baseline. Color coding of missense variants represents likelihood of the variant leading to loss of function according to AlphaMissense. ChEx: Chd1 Exit-side binding domain, DBD: DNA-binding domain, CHCT: CHD C-terminal domain. (B) The distribution of missense and LOF variants in both sexes. (C) Key phenotypic aspects of individuals carrying missense variants predicted to cause loss of protein function and individuals carrying loss of function variants. (D) Phenotypic scores for females and males with curated missense and LOF variants (male, n = 24, female, n = 12). (E) Expected interactions for human CHD1 R618, based on a yeast Chd1-nucleosome structure^32^, which would be potentially disrupted by the missense variant p.R618Q. (F) A schematic of the *in vitro* nucleosome remodeling assay. (G) A representative image from three biological replicates of the chromatin remodeling assay. The CHD1-WT and CHD1-R618Q proteins are denoted by “+” and “-” symbols. The upper band ∼220bp represents the uncut nucleosome-wrapped DNA substrate. The lower band at ∼180bp represents remodeled DpnII-digested DNA-nucleosome substrate. The negative control comprised the DNA alone, with no DpnII restriction site, and the positive control comprised DNA alone with the DpnII restriction site. DpnII was added to all conditions. (H) An overview of the CHD1 protein based on a yeast Chd1-nucleosome structure^32^, with highlighted residues harboring missense variants. \**p*< 0.05, Welch’s two-tailed t-test, ns = not significant.

Collectively, we identified substantial evidence for 36 variants, observed across 40 individuals, which we used to define the core PILBOS phenotype. Key phenotypic characteristics of the 21 individuals carrying putatively disruptive missense variants included developmental delay (86%), speech apraxia/speech delay (81%), and autism spectrum disorder (67%). Two of these individuals were too young to be assessed for intellectual disability, which was present in 68% of the individuals for whom cognitive assessment was possible. For the 19 patients carrying truncating LOF variants, the key phenotypic aspects were speech apraxia/speech delay (95%), autism spectrum disorder (74%), developmental delay (58%) and intellectual disability (44%, with one individual from this group being too young for assessment) (**Figure 1C, Supplementary Tables 1&2**). In order to investigate this further, we scored the probands with curated variants according to the number and severity of core phenotypes. When grouping individuals with missense and LOF variants, females exhibited significantly higher phenotypic scores than males (**Fig. 1D**, *p* = 0.0440, Welch’s t-test), suggesting that females may be more sensitive to *CHD1* variants than males. Together, these data demonstrate that males are overrepresented in the context of PILBOS, whereas females tend to exhibit more severe phenotypic manifestations, suggesting that male sex gives some protection against *CHD1* variants in humans.

### Loss of function of CHD1 caused by specific missense variants

The present cohort carries a high proportion of missense variants in *CHD1* compared to LOF variants, which were not found in the original study^1^. Our computational prediction suggests that many of these missense variants lead to loss of function (**Supplementary Table 1**). To gather more evidence supporting a loss of function mechanism, we performed functional and predictive analyses. First, we selected one representative variant (P_32, **Supplementary Table 1**), a heterozygous variant leading to an amino acid change from a positively charged arginine to neutral glutamine at position 618 in CHD1 which corresponds to the R616Q in mice. This arginine immediately follows the Walker B DExx Box, also known as helicase Motif II (DEAH<u>R</u>), and is strictly conserved in CHD1 orthologs. Since it directly contacts nucleosomal DNA that is actively distorted by CHD1^32^ (**Fig. 1E**), we expected that this arginine-to-glutamine substitution would likely interfere with CHD1 remodeling activity. To assess this, we used a human *CHD1* cDNA construct with an N-terminal deletion, as the N-terminus has been described to inhibit the DNA-binding and ATPase activity of CHD1^19^. We generated the p.R618Q mutation using site-directed mutagenesis, then produced both the wild type (WT) and mutant protein. Using a recombinant DNA-wrapped nucleosome with a DpnII restriction site, we incubated it with DpnII, ATP and either CHD1-WT or CHD1-R618Q at 37°C for multiple different exposure times (**Fig. 1F-G**). While CHD1-WT was rapidly able to shift the nucleosome along the DNA and allow access for DpnII activity in the presence of ATP, CHD1-R618Q was not (**Fig. 1G**, lower band). Thus, the p.R618Q variant causes deficient CHD1 chromatin remodeling activity. Finally, we analyzed the locations of the other missense variants in high resolution structures of human and yeast CHD1 (overview in **Fig. 1H**). Most variants flagged as severe are themselves highly conserved and/or interact with highly conserved residues and motifs and thus would be expected to interfere with enzymatic function (**Fig. S1A**).

### Characterization of *Chd1^R^*^616^*^Q^* mice

In order to examine whether PILBOS phenotypes are observed in a murine model and explore their malleability, we used CRISPR-Cas9 to generate a mouse model harboring the equivalent of the *CHD1^R^*^618^*^Q^* variant (*Chd1^R^*^616^*^Q^*). Given previous reports of homozygous lethality in a *Chd1* knockout mouse model^11^ but not in a mouse model carrying a hypomorphic variant^33^, we sought to investigate the effects of *Chd1^R^*^616^*^Q^*in homozygosity. Over 5 generations of heterozygote-heterozygote breeding, we never observed a homozygous mouse out of a total of 96 pups from 18 litters. Furthermore, we observed decreased Mendelian ratios for mice carrying the variant in heterozygosity: given the number of 39 *Chd1^+/+^* mice, we would expect 78 heterozygotes according to normal Mendelian ratios, but instead observed 57 (**Supplementary Table 3**, p = 2.72 × 10^-9^, Fisher’s exact test). These data suggest functional importance of this variant on fitness either at conception or *in utero*. We next assessed differences in gene expression in cells with the *Chd1^R^*^616^*^Q^* variant compared to cells from *Chd1^+/+^* littermates. Using RNA extracted from the cortex of *Chd1^R^*^616^*^Q/+^*mice, we performed RT-qPCR and observed no significant difference in *Chd1* mRNA levels compared to WT littermates (**Fig. S1B**). We then derived, cultured and extracted protein from mouse embryonic fibroblasts (MEFs) from both genotypes. A western blot demonstrated no significant difference in total CHD1 protein quantity (**Fig. S1C-D**). Thus, although the R616Q variant affects chromatin remodeling activity, it does not appear to do so by reducing the stability of the transcript or the protein.

As growth retardation is a common feature of PILBOS, we examined the weight of the mice over time, starting at postnatal day 21. While the males did not show any significant difference in weight over time (**Fig. 2A**), the females showed a significant weight deficit at postnatal days 21, 46 and 61 (*p* < 0.05, mixed effects analysis, **Fig. 2B**). Using linear regression analysis, we observed a significant difference in the elevation of the regression line between *Chd1^R^*^616^*^Q/+^* mice and WT littermates for females (*p* = 0.0004) but not males (*p* = 0.4883). Since hypotonia is a common PILBOS phenotype, we assessed this feature in the mouse model. Indeed, female *Chd1^R^*^616^*^Q/+^*mice displayed an impaired righting reflex at postnatal day 6 (*p* = 0.0181, Welch’s t-test), which was not observed in males (**Fig. 2C**). In contrast, when using the hindlimb suspension test, which measures strength and endurance, we did not observe a difference in either females (**Fig. S2A**) or males (**Fig. S2B**). To further assess behavioral features in our mouse model, we performed an open field test at 3 months of age, in which the mice were allowed to freely explore an open space for 10 minutes. The female *Chd1^R^*^616^*^Q/+^* mice spent significantly less time in the center of the open field compared to their WT littermates (*p* = 0.0267, Welch’s t-test, **Fig. 2D-E**), a known anxiety-like behavior in mice^34^. Again, no abnormality was observed in male *Chd1^R^*^616^*^Q/+^* mice (**Fig. 2E**). In contrast, there was no difference in the overall distance travelled for either females (**Fig. S2C**) or males (**Fig. S2D**). Intellectual disability is another common PILBOS feature, however female *Chd1^R^*^616^*^Q/+^* mice did not display memory deficits in a Morris water maze test, an established test of spatial learning and memory^35^ (**Fig S2E-H**). However, they travelled less distance (*p* = 0.0119, Welch’s t-test, **Fig. S2I**) and swam with decreased velocity in the water maze (*p* = 0.0148, Welch’s t-test, **Fig. S2J**) compared to WT littermates, concordant with a potential adult motor deficit. Given that speech apraxia and autism are also frequent features of PILBOS, we assessed the ultrasonic vocalizations (UVs) of pups, a measure which can be used to assess differences in social communication, an autism-like phenotype^36^. Measuring every two days from postnatal day 3-9, we did not observe any major differences (**Fig. S2K-L**). Since abnormal eye morphology appears as one of the most significant phenotypes identified in *Chd1* KO mice in data from the International Mouse Phenotyping Consortium (www.mousephenotype.org)^37^, we also examined the structure and electrophysiological responses of the posterior chamber (retina, retinal pigment epithelium and choroid) of the eyes of *Chd1^R^*^616^*^Q/+^* mice. However, we observed no significant differences from their WT littermates (**Fig. S2M-N**). Taken together, *Chd1^R^*^616^*^Q/+^* mice display female-limited weight, motor and behavioral phenotypes.

**Figure 2:**
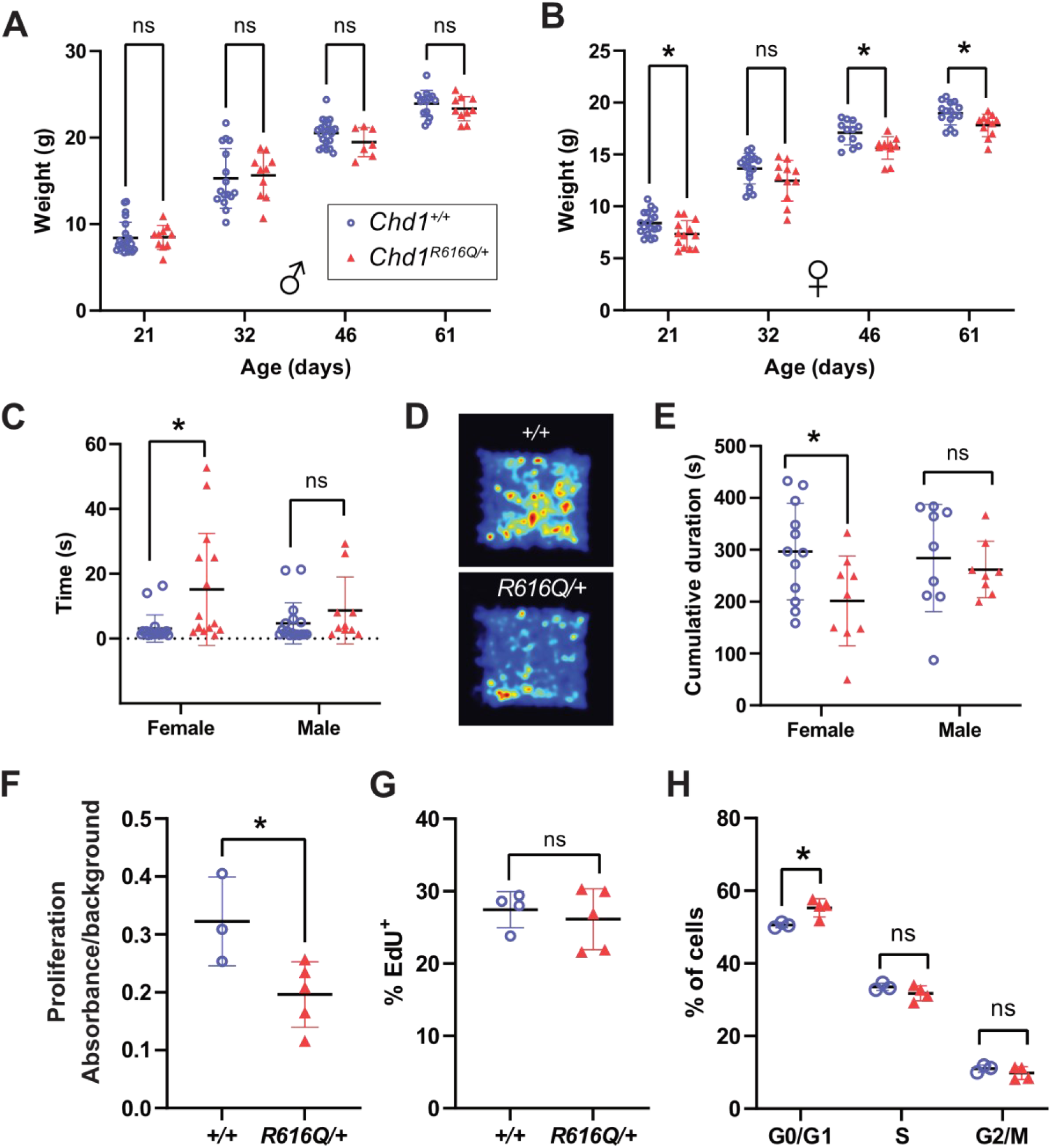
*Chd1^R^*^616^*^Q/+^* mice exhibit female-limited growth, motor and behavioral phenotypes. (A) Weight of male *Chd1^R^*^616^*^Q/+^* mice (*n* = 7-10) and WT littermates (*n* = 14-23). (B) Weight of female *Chd1^R^*^616^*^Q/+^* mice (*n* = 11-13) and WT littermates (*n* = 12-18). (C) Righting reflex times in female (WT: *n* = 20, *R616Q/+*: *n* = 15) and male (WT: *n* = 18, *R616Q/+*: *n* = 10) *Chd1^R^*^616^*^Q/+^* mice and WT littermates at postnatal day 6. (D) A representative heatmap depicting the arena area covered by female mice of both genotypes during a 10-minute open field test. (E) Cumulative duration of time spent in the center area of the open field for female (WT: *n* = 12, *R616Q/+*: *n* = 9) and male (WT: *n* = 9, *R616Q/+*: *n* = 8) *Chd1^R^*^616^*^Q/+^* mice and WT littermates during a 10-minute test. (F) Proliferation of female NPCs in culture as measured by absorbance over background in a MTT assay (WT: *n* = 3, *R616Q/+*: *n* = 5). (G) Flow cytometry measurements of EdU incorporation into female NPC DNA over a 2h period in culture (WT: *n* = 3, *R616Q/+*: *n* = 5). (H) DNA content in female NPCs as measured by flow cytometry following DAPI staining of ethanol-permeabilized cells (WT: *n* = 3, *R616Q/+*: *n* = 4). *p< 0.05, ns: not significant.

### *Chd1^R^*^616^*^Q/+^* mice demonstrate cellular phenotypes in neuronal progenitors

As key PILBOS phenotypes are linked to neuronal function, we next investigated cellular phenotypes in cortical neuronal progenitor cells (NPCs) derived from E18.5 female mouse embryos. Using the 3-(4,5-dimethylthiazol-2-yl)-2,5-diphenyltetrazolium bromide (MTT) assay as a measure of proliferation, we observed significantly decreased proliferation in the *Chd1^R^*^616^*^Q/+^*NPCs compared to NPCs from WT littermates (*p* = 0.0354, Student’s two-tailed t-test, **Fig. 2F**). We then assessed whether the proliferation defect was due to differences in the speed of progression through S-phase by using a 5-ethynyl-2’-deoxyuridine (EdU) assay. Using a 2-hour (h) pulse, we did not observe a difference in the short-term rate of replication (**Fig. 2G**). However, when we assessed the cellular DNA content using DAPI staining, we observed a higher proportion of *Chd1^R^*^616^*^Q/+^* NPCs in the G0/G1 phase of the cell cycle, suggesting a possible delayed entry to S phase, a phenomenon known to occur during neuronal differentiation^38^ (*p* = 0.025, Student’s two-tailed t-test, **Fig. 2H**). *Chd1^R^*^616^*^Q/+^* NPCs did not show any change in the proportion of apoptotic cells as measured by Annexin V assay (**Fig. S2O**). In summary, the R616Q mutation causes decreased proliferation likely through lengthening of the G1 phase of the cell cycle in NPCs, a possible mechanism for the cellular phenotypes observed with disruption of CHD1 in NPCs.

In order to further examine the functional impact of the p.R616Q variant, we performed bulk RNA-sequencing (RNAseq) using female NPCs derived from both genotypes. Modest transcriptomic changes were observed, with 44 significantly differentially expressed genes identified between WT and *Chd1^R^*^616^*^Q/+^* (**Fig. S3A**, **Supplementary Table 4**). In order to not miss any relevant changes, we expanded to a list of genes with the top 10% most significant p-values (*n* = 1809; *p* ranging from 3.33×10^−15^ to 0.08).

Using this list, we performed overrepresentation analysis and observed “microfibril”, “neuron-to-neuron synapse” and “centrosome” amongst the most significantly enriched cellular component terms (**Fig. S3B**). Next, using molecular pathway enrichment, we observed “oxytocin signaling pathway” amongst the significantly overrepresented pathways (**Fig. 3A**). This finding is particularly noteworthy as oxytocin is known to exhibit sexually dimorphic effects on weight and anxiety^39–44^. Genes encoding the oxytocin receptor (*Oxtr*) and downstream effectors including *Gnaq*, *Ptgs2*, *Prkcg*, *Rhoa* and *Eef2k* were upregulated in the *Chd1^R^*^616^*^Q/+^* cells compared to wildtype (**Fig. 3B**). To investigate the functional implications of this observation, we conducted an enzyme-linked immunoassay (ELISA) on protein extracted from hypothalamic tissue of 3-month-old female mice and observed significantly elevated levels of oxytocin in the *Chd1^R^*^616^*^Q/+^* mice compared to WT littermates (*p* = 0.0337, Welch’s t-test, **Fig. 3C**). These results suggest that even genes that did not meet the more stringent significance threshold may still hold biological relevance, justifying our approach of considering genes in the top 10% of p-values.

**Figure 3:**
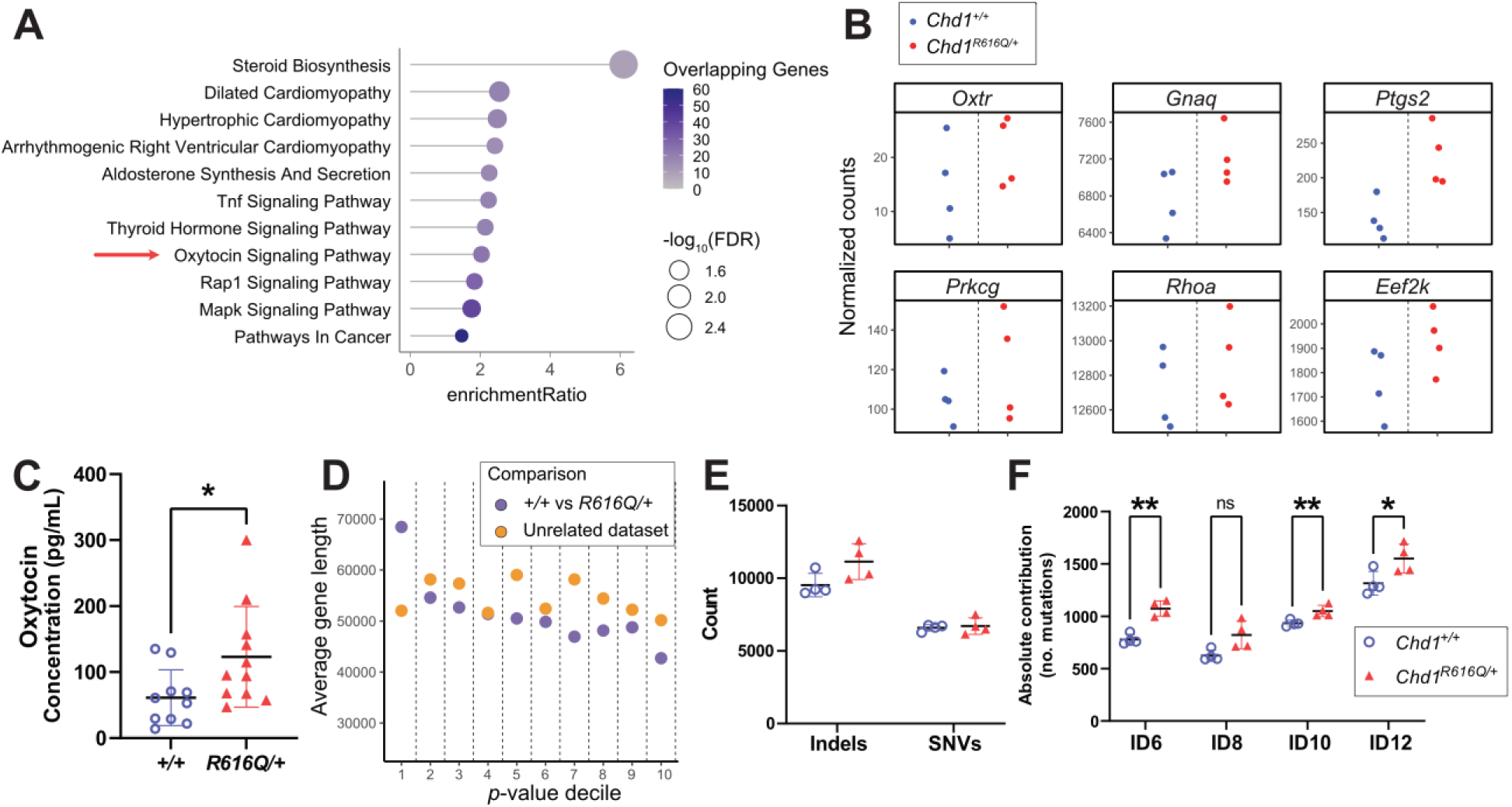
Gene expression in *Chd1^R^*^616^*^Q/+^* NPCs shows changes in oxytocin signaling, enrichment of long genes and a distinct mutational signature. (A) KEGG pathway gene ontology overrepresentation analysis for the comparison of gene expression results from *Chd1^+/+^* vs *Chd1^R^*^616^*^Q/+^*. (B) Normalized RNAseq counts for genes involved in the oxytocin signaling pathway in *Chd1^+/+^* and *Chd1^R^*^616^*^Q/+^* NPCs, selected from the gene ontology overrepresentation analysis. (C) Oxytocin concentration in hypothalamic lysates from 3-month-old female *Chd1^R^*^616^*^Q/+^* mice and WT littermates as measured by ELISA (WT: *n* = 10, *R616Q/+*: *n* = 11). (D) Average gene lengths for each *p*-value decile, with 1 being the most significant *p*-values for the WT vs *Chd1^R^*^616^*^Q/+^* comparison and an unrelated wild-type NPC dataset. (E) Counts of indels and SNVs in RNAseq data. (F) Counts of mutations associated with COSMIC indel mutational signatures. \**p*< 0.05, \*\**p* < 0.01.

As CHD1 is known to play roles in DNA damage repair^17–20^, and long genes are more susceptible to DNA damage which can affect their transcription^45^, we next assessed the gene-lengths of detected genes in our RNAseq datasets ranked according to *p*-value. Here we observed that the genes with the top 10% most significant p-values (decile 1), were significantly longer than all other detected genes (*p* = 2.973 × 10^-6^, Welch’s t-test, **Fig. 3D**). In contrast, when looking at an unrelated WT RNAseq dataset also generated from E18.5 WT mouse NPCs^46^, there was no significant difference in gene length for the most significantly differentially expressed genes (*p* = 0.3084, Welch’s t-test **Fig. 3D**). Next, we assessed the number of indels and single nucleotide variants (SNVs) in the RNAseq data. We observed a slight but non-significant increase in the number of indels in the *Chd1^R^*^616^*^Q/+^* NPCs compared to WT (*p* = 0.07, Welch’s t-test), whereas the number of SNVs was unchanged (**Fig. 3E**). Further analysis of the COSMIC mutational signatures^47^ associated with the indels revealed the signature “ID1” as most enriched, which consists primarily of T-base insertions. Given that this pattern could reflect poly(A) tails, which are expected in RNA-seq data, we excluded ID1 from further analysis to avoid potential confounding. Among the remaining signatures, “ID6”, “ID8”, “ID10” and “ID12” were notably enriched and three were significantly increased in *Chd1^R^*^616^*^Q/+^* NPCs relative to WT (ID6: *p* = 0.0008, ID8: *p* = 0.0517, ID10: *p* = 0.0144, ID12: *p* = 0.0399, Welch’s t-test, **Fig. 3F**). While ID10 and ID12 are not associated with any specific etiology, ID6 is associated with defective homologous recombination DNA damage repair and ID8 has a proposed etiology linked to repair of double-strand breaks by non-homologous end-joining (NHEJ). These findings suggest that *Chd1^R^*^616^*^Q/+^* NPCs may be deficient in homologous recombination activity, which is known to be dependent on CHD1^17,19,20^, and instead rely more heavily on the error-prone NHEJ pathway, potentially implicating DNA damage in the etiology of the PILBOS phenotype.

### Androgen exposure rescues *Chd1^R^*^616^*^Q/+^* transcriptional abnormalities

Previous studies have shown direct protein interactions between CHD1 and the AR^24,25^. We hypothesized that the greater androgen exposure that naturally occurs in males could be a potential mechanism mediating the male protective effect in our *Chd1^R^*^616^*^Q/+^* mice and human clinical cohort. To test this, we performed RNAseq using NPCs treated with 10nM dihydrotestosterone (DHT) or with DMSO vehicle control (**Supplementary Table 5**, **Fig. S4A**). As before, we collected the genes with the top 10% most significant *p*-values from the comparison of DHT vs DMSO control treatment in *Chd1^R^*^616^*^Q/+^* cells (*n* = 1960, *p* ranging from 6.66 × 10^-16^ to 0.08). The most significantly enriched cellular component gene ontology terms in this list also included “neuron-to-neuron synapse” and “centrosome”, as well as “cytosolic ribosome” (**Fig. S4B**). KEGG pathway analysis also revealed enrichment of the term “ribosome”, which may imply differences in regulation of protein synthesis, a known effect of androgens^48–50^ (**Fig. S4C**). Interestingly, for several genes whose expression was the most significantly changed in the *Chd1^R^*^616^*^Q/+^* cells compared to WT, DHT treatment of the *Chd1^R^*^616^*^Q/+^* cells restored the expression towards baseline WT levels (**Fig. 4A**). We next compared these two lists (WT vs *Chd1^R^*^616^*^Q/+^* and control vs DHT treatment in *Chd1^R^*^616^*^Q/+^*cells) and observed a highly significant overlap of 425 genes (*p* = 4.1 × 10^-73^, OR = 3.3, Fisher’s exact test, **Fig. 4B**). Among the most significantly enriched gene ontology terms for this overlapping list of genes were “glutaminergic synapse”, “cytoskeletal part” and “neuron part”, with genes including *Agrn*, *Bcan*, *Syt1, Dab1 and Ntrk3,* suggesting changes in neuronal function and differentiation (**Fig. 4C**). We then sought to understand the relationship between the changes caused by the R616Q variant and DHT. When we plotted the log_2_ fold change for the overlapping genes from each comparison against one another, we observed a strong negative correlation (*r* = -0.916, *p* < 2.2 × 10^-16^, **Fig. 4D**). This indicates that genes which were over-expressed in *Chd1^R^*^616^*^Q/+^*cells exhibit lower expression following DHT treatment and vice versa, suggesting that DHT treatment reverses the transcriptional dysregulation caused by the R616Q variant in NPCs. In contrast, when we looked at the lists of genes with the 10% least significant *p*-values from the two comparisons, there was an overlap of 181 genes, which was not statistically significant. Furthermore, when we plotted the log_2_ fold change for these genes, we did not observe a correlation (*r* = 0.102, *p* = 0.1731, **Fig. 4E**), demonstrating that this effect is specific to the most highly differentially expressed genes. As testosterone is converted to estradiol by the aromatase cytochrome P450 enzyme^51^, we examined the expression of this gene (*Cyp19a1*) and the estrogen receptor genes in the NPCs and observed relatively low levels of expression compared to the androgen receptor (**Fig. S4D**). Together with the fact that the non-aromatizable androgen, DHT restores normal expression in mutant cells, this suggests the effects of testosterone treatment may be more likely to be driven by AR activation rather than the estrogen receptor, at least in neural progenitor cells.

**Figure 4:**
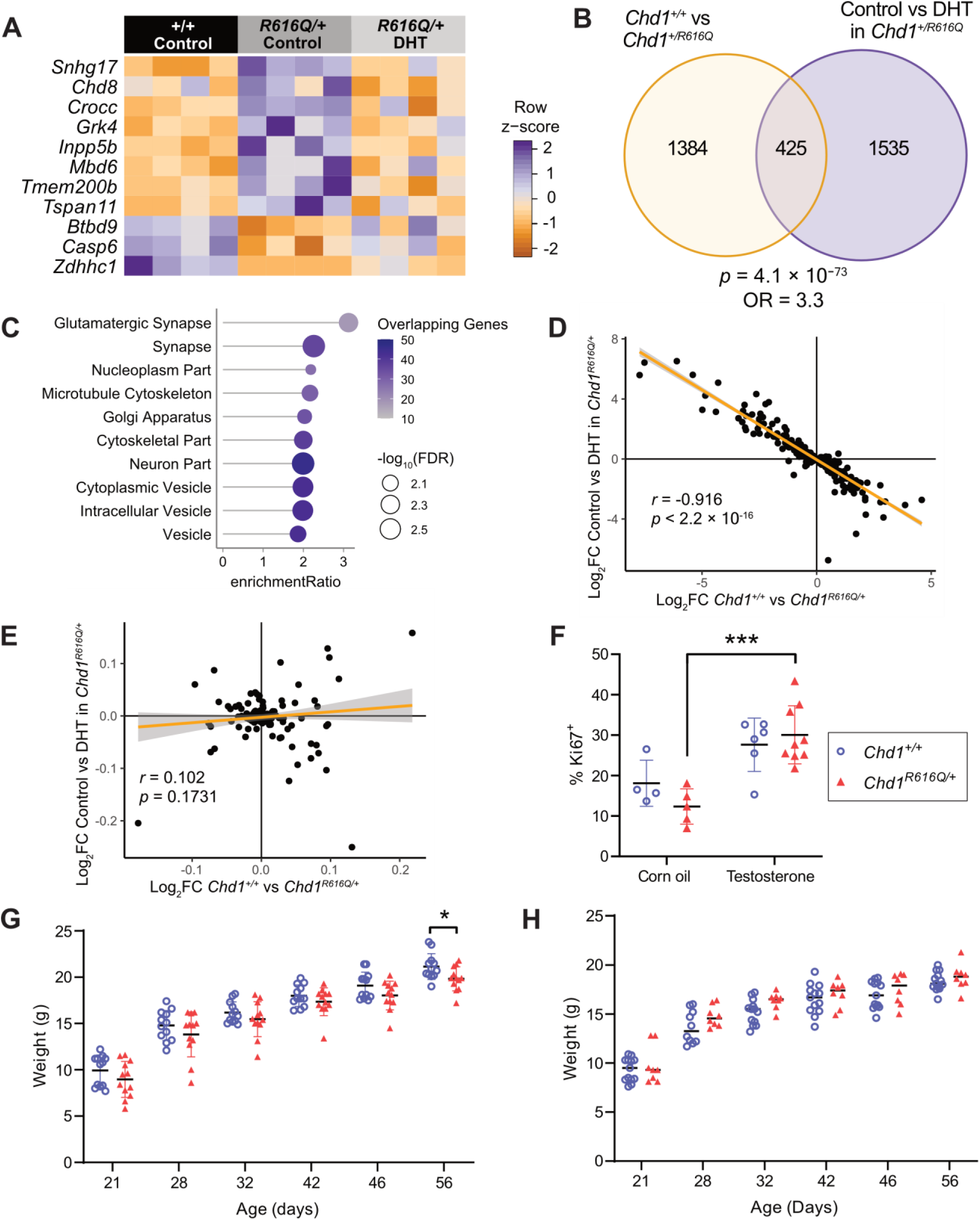
Androgens rescue *Chd1^R^*^616^*^Q/+^* transcriptomic and *in vivo* phenotypes. (A) A heatmap depicting mRNA expression in NPCs, either WT or *Chd1^R^*^616^*^Q/+^* treated with DMSO vehicle control, or *Chd1^R^*^616^*^Q/+^* treated with 10nM DHT for 6h. (B) A Venn diagram displaying overlapping genes with the top 10% most significant p-values from the comparison between *Chd1^+/+^* vs *Chd1^+/R^*^616^*^Q^* and DMSO vehicle control vs DHT in *Chd1^+/R^*^616^*^Q^*NPCs. The overlap was tested using Fisher’s exact test with a background of 19980 genes (number of protein coding genes in GENCODE hg38). (C) Overrepresentation analysis of overlapping gene list from B. (D) Plot of log_2_ fold changes for the genes with the 10% lowest p-values from the overlap in B. (E) Plot of log_2_ fold changes for the overlapping genes with the 10% highest p-values in the “WT vs *Chd1^R^*^616^*^Q/+^*” and “DMSO vs DHT in *Chd1^R^*^616^*^Q/+^*” comparisons. (F) Percent Ki-67^+^ cells in the telencephalon of embryonic day 18.5 embryos, as measured by antibody staining and flow cytometry (WT-corn oil: *n* = 4, *R616Q/+*-corn oil: *n* = 5, WT-testosterone: *n* = 6, *R616Q/+*-testosterone: *n* = 9). Embryos were obtained from 3-4 dams for each treatment. (G) Weight measurements over time in orchiectomized male *Chd1^R^*^616^*^Q/+^* mice and WT littermates (*n* = 12 per genotype). (H) Weight measurements over time in female mice with a testosterone implant placed under skin at P15 (WT: *n* = 10-13, *R616Q/+*: *n* = 8). *p < 0.05, ***p < 0.001.

### Androgens ameliorate *in vivo Chd1^R^*^616^*^Q/+^* phenotypes

Given the reversal of *Chd1^R^*^616^*^Q/+^* -driven transcriptional abnormalities with androgen exposure in NPCs *in vitro*, we next sought to investigate if androgen exposure could rescue *in vivo* phenotypes in our mouse model. As we had previously observed decreased proliferation of NPCs from the *Chd1^R^*^616^*^Q/+^*model (**Fig. 2F**), we sought to test whether testosterone treatment would ameliorate this defect. Indeed, 10 days of testosterone injection of pregnant dams led to increased proliferation in the telencephalon of *Chd1^R^*^616^*^Q/+^*embryos at E18.5 (*p* = 0.0001, Welch’s t-test, **Fig. 4F**). We hypothesized that if androgens were protecting males from phenotypes driven by CHD1-deficiency, then removal of androgens might unmask these same phenotypes in *Chd1^R^*^616^*^Q/+^* males. To test this, we performed orchiectomy of both WT and *Chd1^R^*^616^*^Q/+^* male mice at postnatal day 15 (P15) and then followed their weight prospectively.

Although there was no significant weight difference between the genotypes prior to orchiectomy (**Fig. S4E**), after the P15 procedure a weight deficit emerged in the orchiectomized *Chd1^R^*^616^*^Q/+^* mice compared to their orchiectomized WT littermates at P56 (*p* = 0.0255, mixed effects analysis, **Fig. 4G**). Using linear regression analysis, we also identified a significant difference in the elevation of the regression line for *Chd1^R^*^616^*^Q/+^* males compared to their WT littermates (*p* = 0.0037). Next, to test if the female weight deficit could be rescued through androgen treatment, we surgically implanted slow-release testosterone silastic implants under the skin of both WT and *Chd1^R^*^616^*^Q/+^* female mice at P15 (**Fig. S4F**). Following their weight over time, we observed amelioration of the previously observed weight deficit in the *Chd1^R^*^616^*^Q/+^* females compared to their WT littermates, with no significant decrease in their weights (mixed effects analysis, **Fig. 4H**). Furthermore, using linear regression analysis, we observed less difference in the elevation of the regression line for *Chd1^R^*^616^*^Q/+^*females compared to their WT littermates (*p* = 0.0254, previously 0.0004). We also tested if orchiectomy would uncover the female-limited anxiety-like behavior in males using an open field test. However, there was no difference in the time spent in the center of the open field following orchiectomy (**Fig. S4G**), suggesting that the sequence of events underlying postnatal anxiety-like behavior may occur prior to P15 (time of orchiectomy). Taken together, our data demonstrate a protective effect of androgens against weight and neuronal proliferation deficits in *Chd1^R^*^616^*^Q/+^* mice.

### Rare *CHD1* missense alleles are overrepresented in healthy males

Given the overrepresentation of males and the increased phenotypic scores of females in our PILBOS cohort, along with the protective effect of androgens in our mouse model, we sought to explore if this could be attributed to a general sex-specific susceptibility to *CHD1* variants. In exome sequencing data from the Genome Aggregation Database (gnomAD, v4.1.0)^30^, which mostly excludes individuals with pediatric disease, we assessed the frequency of alternate alleles in *CHD1* in female (XX) and male (XY) populations. As previously reported^1^, *CHD1* is highly intolerant to loss of function variants (pLI = 1) so there are few LOF variants to evaluate. Considering all missense variants, we decided to filter out those with a minor allele frequency (MAF) of ≥ 1% as these are much less likely to have functional effects.

Using the remaining rare missense variants, we calculated the aggregate sum of alternate allele counts (AC) divided by the aggregate sum of total allele number (AN) for each sex and compared these frequencies (AF) between the sexes using a two-proportion z-test. We observed a significant overrepresentation of missense alleles in males (AF = 1.35 × 10^-5^) compared to females (AF = 1.24 × 10^-5^) (*p* = 3.65 × 10^-10^, χ² = 39.29, 95% CI = 5.73 × 10^-7^ -1.37 × 10^-6^, **Fig. 5A**). To test the validity of our findings, we next repeated this analysis in whole genome sequencing (WGS) data from the UK biobank (UKB)^52,53^. Here, rare missense variants in *CHD1* were also overrepresented in males (AF = 1.53 × 10^-5^) compared to females (AF = 1.47 × 10^-5^) (*p* = 0.019, χ² = 5.48, 95% CI = 4.46 × 10^-8^ - 5.10 × 10^-7^, **Fig. 5A**). Together, these data suggest that healthy males (i.e., those without rare pediatric conditions) are more likely than females to carry rare missense variants in *CHD1*, indicating a sex-differential susceptibility to these variants. In the All of Us dataset^54^, sex at birth was not associated with being a carrier of at least one rare (MAF < 1%) missense variant in *CHD1* (*p* = 0.15, two-sided z-test, logistic regression, Methods), potentially reflecting differences in recruitment practices, demographic composition, or variant calling pipelines across cohorts.

**Figure 5:**
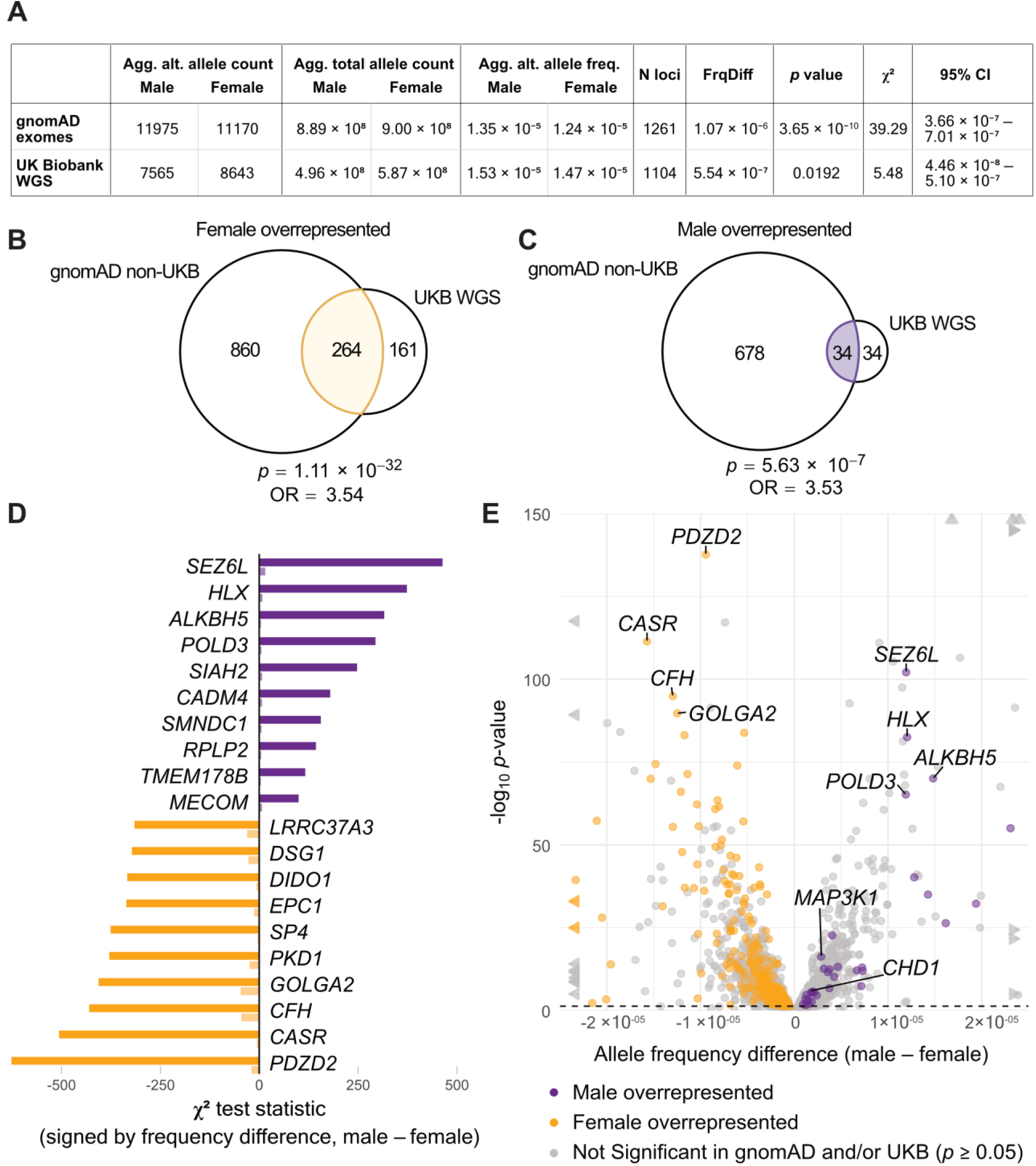
Sex-bias of rare missense variants in *CHD1* and other highly constrained genes. (A) Comparison of aggregated alternate allele frequencies of rare (MAF < 1%) missense variants across males and females in the *CHD1* gene in the total gnomAD exomes and UK Biobank WGS datasets. The two-proportions z-test was used to compare frequencies between sexes. The overlap between genes with nominal (B) female and (C) male overrepresentation (p < 0.05) of rare (MAF < 1%) missense variants for highly constrained (pLI > 0.9) autosomal genes in the gnomAD non-UKB exomes and UKB WGS datasets. The *p* values and odds ratios for the overlaps were calculated using Fisher’s exact test using a background of 19980 genes (number of protein coding genes in GENCODE hg38). (D) Chi-squared test statistic multiplied by the sign of the aggregated allele frequency differences for the top 10 genes from the overlapping gene lists. The sign corresponds to the direction of the aggregated allele frequency difference (male – female). (E) Sex differences in rare (MAF < 1%) missense variant frequencies for highly constrained (pLI > 0.9) autosomal genes. Each dot represents a gene, showing the aggregated alternate allele frequency difference (male – female) against the -log_10_ *p*-value from the gnomAD non-UKB exomes dataset. Statistics were computed using two-proportions z-test. Arrows represent genes which exceed the plot range. The dashed line represents the *p* = 0.05 threshold for the gnomAD dataset analysis. Colored genes are those with *p*-values < 0.05 in both the gnomAD non-UKB exomes and UKB WGS datasets.

### Autosomal genes show sex-biased allele frequencies in healthy populations

Next, we extended our analysis to all autosomal genes with a probability of loss of function intolerance (pLI) score of > 0.9, i.e. highly constrained genes. Using the gnomAD exome sequencing database excluding UK biobank samples, out of a total of 3140 genes, we observed 663 genes with a significant overrepresentation of rare (MAF < 1 %) missense variants in males and 1060 genes with female overrepresentation after Benjamini-Hochberg multiple-testing correction (*p*-adj < 0.05, **Supplementary Table 6**). To assess the impact of variant frequencies on sex-specific overrepresentation, we next compared gene lists obtained using lower MAF thresholds (< 0.5% and < 0.1%) and observed similar results (**Fig. S5A-D**), suggesting that the observed sex bias is largely driven by very rare variants.

We repeated the sex overrepresentation analysis for rare missense variants (MAF < 1%) in the UKB WGS dataset, and also in the total gnomAD exomes dataset and the UK Biobank-only exomes dataset in gnomAD, observing positive correlations between frequency difference values (male AF – female AF, **Fig. S5E-G**). Using the UKB WGS dataset analysis, we identified 6 genes with male overrepresentation and 124 genes with female overrepresentation after Benjamini-Hochberg multiple-testing correction (*p*-adj < 0.05, **Supplementary Table 7**). To expand upon the analysis, we then examined all genes with *p*-values < 0.05 from the gnomAD non-UKB and UKB WGS datasets. Of the 425 genes with nominally significant female overrepresentation in the UKB WGS dataset, 264 genes overlapped with nominally significant female-biased genes in the non-UKB gnomAD dataset (*p* = 1.11 × 10^-32^, OR = 3.54, **Fig. 5B**). Of the 68 genes with nominally significant male overrepresentation in UKB, 34 overlapped with genes with male overrepresentation in gnomAD (*p* = 5.63 × 10^-7^, OR = 3.53, **Fig. 5C**). This list included *CHD1* and *MAP3K1*, where gain-of-function variants in *MAP3K1* are known to cause gonadal dysgenesis in 46,XY individuals^55,56^, while loss-of-function variants impair female reproductive development^57^. The top 10 overlapping genes with male overrepresentation included *SEZ6L*, *HLX* and *ALKBH5*, while female-biased genes included *PDZD2*, *CASR* and *CFH* (**Fig. 5D**). The results were similar using lower MAF thresholds (< 0.5% and < 0.1%, **Fig. S5H-K**).

Overall, fewer genes exhibited sex bias in the UKB WGS dataset compared to the gnomAD non-UKB exomes dataset, with the most pronounced difference observed among genes with male overrepresentation. Notably, the UKB-only exome data from gnomAD showed a similar number of genes with male or female sex bias as the UKB WGS dataset using the MAF < 1% threshold (**Fig. S5L-M**).

Differences between the gnomAD and UKB datasets may therefore reflect disparities in cohort compositions. The UK Biobank is known to exhibit ascertainment bias toward healthier individuals with higher income and educational attainment^58,59^, 88.26% of whom self-identify as British/White.^60^. The gnomAD non-UKB v4 exomes dataset is drawn from case-control studies of adult-onset disorders as well as diverse biobanks from around the world and includes a higher proportion of individuals with non-European genetic ancestries^30^ (35.87%, https://gnomad.broadinstitute.org/stats). Thus, using two independent population datasets, we have identified 298 genes that display sex-biased allele frequencies of rare missense variants (**Fig. 5E**, **Supplementary Table 8**).

### Neurodevelopmental functions are enriched in autosomal sex-biased genes

A subset of our sex-biased genes may be true examples of disorders with male or female protective effects. Any sexual dimorphisms may however hinder the ability to define them as disease-causing genes. When we assessed the overlap of our sex-biased gene list with a published list of genes with sex-biased expression in human organs^61^, *CHD1* (overrepresented in males), *VPS54, TTC7B* and *LIMK1* (overrepresented in females) appeared in both lists, with differential expression in brain tissue.

We then looked deeper into the molecular functions of the genes in each list by performing overrepresentation analysis. Genes with female overrepresentation in both gnomAD and UKB were significantly enriched for molecular function gene ontology terms relating to “vascular endothelial growth factor receptor activity”, “histone deacetylase activity”, “transmembrane receptor protein kinase activity” and transcriptional regulation (**Fig. S6A**, FDR < 0.1), and for Reactome pathway terms “axon guidance” and “nervous system development” (**Fig. 6A**, FDR < 0.1). In contrast, while genes with male overrepresentation were not significantly enriched for molecular function gene ontology terms, they were significantly enriched for the Reactome pathway term “netrin mediated repulsion signals”, “netrin-1 signaling” and “developmental biology” (**Fig. 6B**, FDR < 0.1).

**Figure 6:**
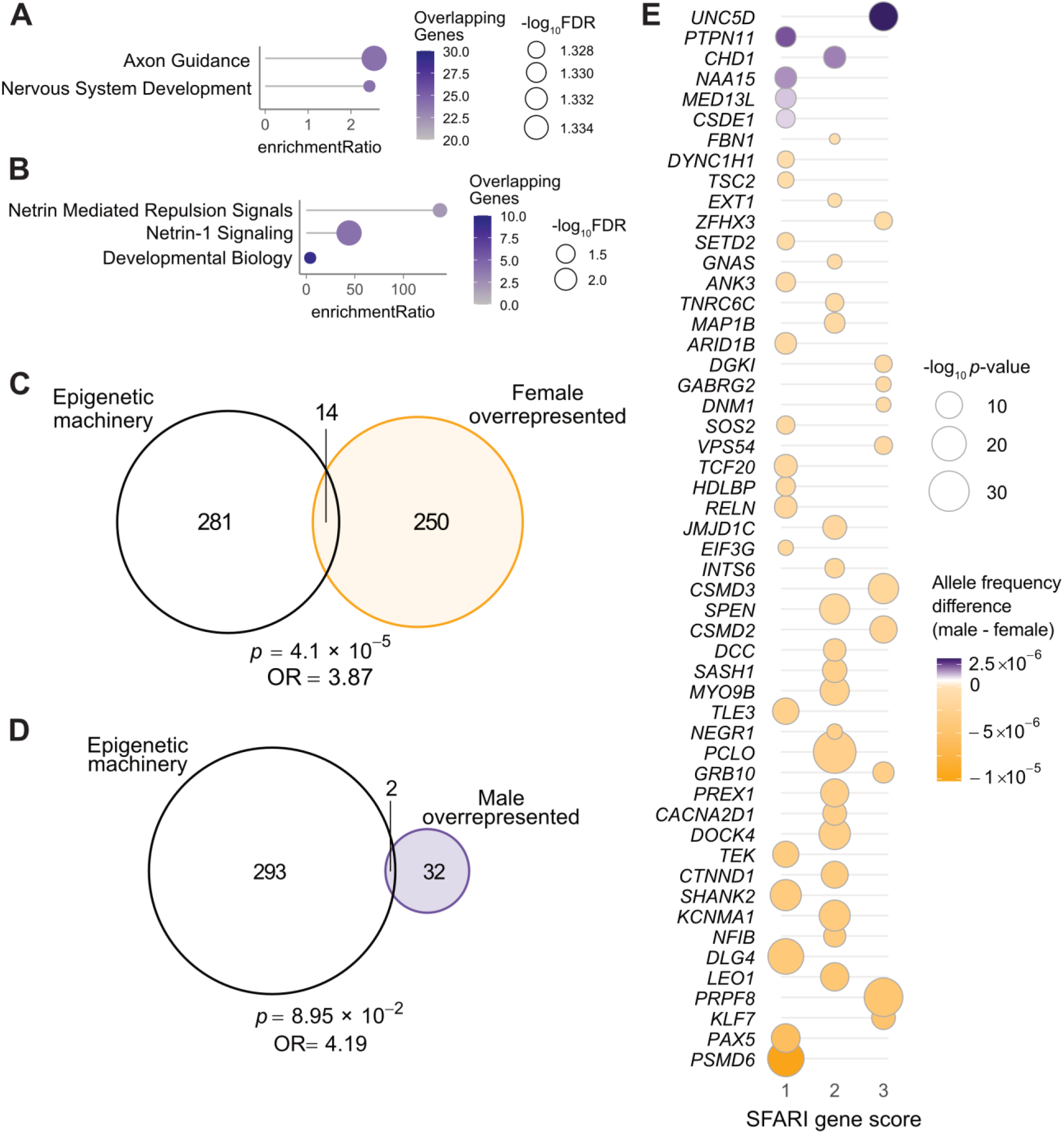
Overrepresentation of neurodevelopmental, epigenetic and autism-related genes among those showing sex bias in variant frequencies

Given that terms relating to histone deacetylation, transcriptional regulation and neurological development were amongst the most significantly enriched for the list of genes with female overrepresentation, we next examined overlap with the epigenetic machinery (EM). EM genes are a known class of genes that are highly intolerant to loss of function variation and frequently linked to NDD, with *CHD1* being a part of this group^62^. Using a pre-defined EM gene list^62^, we observed a significant overlap of 14 EM genes with female overrepresentation in gnomAD and UKB including *DNMT1*, *HDAC3* and *JMJD1C* (*p* = 4.1 × 10^-5^, OR = 3.87, **Fig. 6C**), and a non-significant overlap of 2 EM genes with male overrepresentation, *CHD1* and *SMNDC1* (*p* > 0.05, **Fig. 6D**, epigenetic function denoted in **Supplementary Table 8**). Notably, the *SMNDC1* transcript is a known target of repression by the fragile X mental retardation protein (FMRP)^63,64^.

As autism is an NDD with known sex-bias, we next assessed the overlap of the genes we identified with female or male overrepresentation with the SFARI Gene autism candidate list^65^. We identified a significant overlap of 48 SFARI genes with female overrepresentation (*p* = 5.1 × 10^-12^, OR = 3.57, **Fig. S6B**) and a significant overlap of 6 SFARI genes with male overrepresentation (*p* = 0.015, OR = 3.36, **Fig. S6C**). Among these, we observed 2 with a “Syndromic” classification but no score, 20 with a SFARI gene score of 1 (high confidence), 21 with score 2 (strong candidate), and 11 with score 3 (suggestive evidence) (**Fig. 6E**, **Supplementary Table 8**). Although only 6 of the genes with male overrepresentation overlapped with SFARI autism genes, additional genes are associated with neurodevelopmental and psychiatric disorders including *ADD3*^66^, *FBXW7*^67^, *POLD3*^68^ and *SEZ6L*^69^. These findings highlight a notable enrichment of neurodevelopment-associated genes with significant differences in the frequency of rare missense variants between males and females, suggesting that genetic factors contributing to neurodevelopment may be particularly prone to sex-biased penetrance.

Overrepresentation analysis of genes found in both gnomAD and UKB with (A) female overrepresentation and (B) male overrepresentation using the reactome pathways gene sets. (C) Overlap of genes that show female overrepresentation in gnomAD and UKB datasets with epigenetic machinery genes. (D) Overlap of genes that show male overrepresentation in gnomAD and UKB datasets with epigenetic machinery genes. The *p* values and odds ratios for the overlaps were calculated using Fisher’s exact test with a background of 19980 genes (number of protein coding genes in GENCODE hg38). (E) Genes with male or female overrepresentation in gnomAD and UKB that overlap with SFARI autism candidate genes. SFARI gene score is shown on the x-axis, 1: High confidence, 2: Strong candidate, or 3: Suggestive evidence. The male - female aggregated alternate allele frequency difference is depicted by the color of the circles and -log_10_ *p*-value is depicted by the size of the circles.

## Discussion

In the present study, we have extended the understanding of PILBOS, highlighting key clinical information from the largest PILBOS cohort assembled to date. We have demonstrated validity of a new PILBOS mouse model, which may serve as a valuable reagent to the research community in the future.

Most importantly we have identified a protective role of androgens against two of the disorder phenotypes, growth retardation and the neuronal proliferation defect in mice, and demonstrated a sex-bias in *CHD1* variants on the human population level. Furthermore, we have identified additional highly constrained genes with a similar sex bias, suggesting that other autosomal Mendelian disorders may show similar sexual dimorphism, and this may be worth further study in murine models and clinical settings.

As no cohort studies of PILBOS have been published since the original description of the disorder, the data presented here will likely prove valuable to clinicians in placing diagnoses and counseling patients as well as to DNA diagnostic laboratories in interpreting variants. Notably, males were overrepresented amongst PILBOS probands and females had more severe phenotypic scores, suggesting that males may have increased tolerance for disruption of the CHD1 protein compared to females. This may be a useful rule that can be further fleshed out as additional individuals are diagnosed.

Our mouse model may be useful in the future to test potential treatments for PILBOS, or to gain deeper insights into the role of CHD1 in development. Although a prior study observed no phenotypic abnormalities in *Chd1^+/-^* mice^11^, this may be caused by not keeping track of sex of individual animals or the use of different assays to evaluate phenotypes. Another possibility is that missense variants in *Chd1* can have more severe dominant-negative effects, but given the paucity of heterozygous nonsense or null *CHD1* variants in the human population and our functional data in this study, this seems less likely. A second study identified spatial memory deficits in a homozygous mouse model with an N-terminal deletion in the gene encoding the CHD1 protein^70^, which contrasts with our findings in the *Chd1^R^*^616^*^Q/+^*mice. This discrepancy may be due to the presence of a functional N-terminus in our model, or the fact that our mice have one fully functional allele. It could also relate to the absence of intellectual disability in the subject from which the original human p.R618Q variant was derived. Potentially, variants with more profound effects on the CHD1 protein may lead to learning deficits in mouse models that are easier to discriminate from WT littermates.

Notably, oxytocin levels were elevated in the *Chd1^R^*^616^*^Q/+^* mouse model. This may suggest abnormal hypothalamic function, which by itself could lead to both weight and anxiety phenotypes, as increased oxytocin is known to reduce weight gain by decreasing food intake, increasing energy expenditure and promoting lipolysis^39–41^, and is also associated with anxiety^42–44^. Oxytocin itself is also known to drive sexually dimorphic circuits in the brain^71^, and its receptor is expressed in a sexually dimorphic manner^72^. *PTEN* variants, a common cause of ASD, also lead to elevated levels of oxytocin in mouse models^73^, and our finding is thus a second example of elevated oxytocin levels in the hypothalamus in the context of ASD. This link opens avenues to study elevated oxytocin as a novel biomarker of a subset of ASD, which may suggest shared pathogenesis and a potential for shared therapeutic strategies.

The protective role of androgens we demonstrate in this study opens a new path for investigation into therapeutic strategies for PILBOS treatment. Indeed, how do androgens protect against CHD1 deficiency? Testosterone is known to increase survival of neurons in the rodent hippocampus^74,75^ and androgen signaling has been reported to promote proliferation in the mouse cortex during development^76^. In human cerebral organoids, androgens promote excitatory neurogenesis^77^ and studies indicate their neuroprotective effects across multiple brain regions, including the rat hypothalamus and visual cortex^78^, as well as in promoting motor neuron survival in injury models^79^. Thus, androgens may be protective against PILBOS through counteracting the neural proliferation deficit and promoting neuronal survival. At the molecular level, the AR may facilitate chromatin remodeling at sites typically regulated by CHD1, potentially compensating for the deficiency in its normal function. Upon androgen stimulation, the AR is known to recruit pioneer factors, which promote chromatin opening and enhance access to target gene loci^80^. Furthermore, AR activation is protective against genomic instability and DNA damage accumulation, driving transcription of non-homologous end-joining pathway genes and the AR and TOP2B are co-localized to genomic breakpoints following androgen stimulation^81^. Interestingly, the effects of CHD1 deficiency on the transcription of long genes mirrors that of another chromatin remodeler, CHD7, whose interaction with TOP2B maintains transcription of long neuronal genes^82^.

Notably, TOP2B is also a CHD1 interaction partner^21^, indicating a partially shared function of the two remodelers. Another possible driver of PILBOS phenotypes could be reduced androgen levels in PILBOS-affected individuals of either sex, a phenotype which is common with CHD7 deficiency^83,84^. In line with this hypothesis, genes involved in the “steroid biosynthesis” pathway were significantly enriched amongst the most significantly altered genes in NPCs from our mouse model, with almost all being downregulated.

It is interesting to speculate that altered expression of many androgen-responsive genes could give rise to the female-specific defects in PILBOS, which are prevented in the majority of males by their naturally higher levels of androgen signaling, bringing the expression of these genes closer to the baseline level. If this effect occurs widely *in utero*, as is the case for neuronal proliferation in our mouse model, it could lead to increased female embryonic lethality, resulting in fewer females with PILBOS diagnoses. Indeed, combining the clinical data with our analysis of the gnomAD and UK Biobank population data, our findings suggest increased embryonic lethality amongst female carriers of pathogenic *CHD1* variants. In males with the most severe defects in CHD1, normal androgen signaling may be insufficient to overcome the induced changes in gene expression, resulting in the development of PILBOS features. A consequence of this line of thought would be that PILBOS features may improve in males upon puberty. However, since our cohort in this report consists almost entirely of young children, we cannot draw conclusions on this matter at present, but it would be of great interest to follow these individuals prospectively and measure hormone levels such as androgens and hypothalamic hormones. This could open up a potential treatment right away by providing testosterone treatment to those males that have the lowest values.

Overrepresentation of a particular sex in a clinical cohort of a genetic disorder can indicate one of two possibilities: greater susceptibility of that sex to variants in a given gene, or protection against them. Traditionally, the former has been assumed, as seen in arguments for male susceptibility in autism being characterized by male overrepresentation. However, population-level data can challenge this assumption. Using PILBOS—a genetic disorder with male overrepresentation—we illustrate that when a sex is overrepresented both in affected individuals and in the healthy population, this pattern may instead reflect a protective effect in that sex. A similar pattern is seen for the gene *PTPN11*, whose variants we observe to be overrepresented amongst males in the healthy population. *PTPN11* is a cause of Noonan syndrome, which is known to exhibit a male-biased sex ratio, both for *de novo* and familial cases^85^. Furthermore, our analysis revealed variants in the gene *HDLBP* to be overrepresented in females. Notably, *HDLBP* resides within the locus for 2q37.3 microdeletion syndrome, a NDD with a female-biased sex ratio^86^.

Conversely, some genes do support the traditional interpretation of increased susceptibility for the sex that is overrepresented in clinical cohorts. For example, *SEZ6L* was the most significant high-confidence male-overrepresented gene in our population dataset analysis. Variants in *SEZ6L* have been associated with bipolar disorder specifically in females^69^, and *Sez6l* knockout mice show female-limited weight differences and motor-coordination deficits^87^. Another example is *NRG1*, which was overrepresented in females in both healthy population datasets and is linked to sex-specific neurological and behavioral phenotypes that are more severe in male mice^88^ and rats^89^. In humans, decreased circulating NRG1 levels correlate with increased severity of early-onset schizophrenia specifically in males^90^ and variants in *NRG1* are associated with elevated schizophrenia risk in males^91^.

One caveat of our analysis is that the same individual could be counted more than once if they have multiple variants in our gene list. However, the effect of this potential bias is likely minimal given the rarity of the variants and the stringent filtering criteria applied.

Overall, the results of our population database analyses are congruent with the concept of a female-protective effect being more common in genetic disorders, with many more genes showing female rather than male overrepresentation. However, the notable number of genes with male overrepresentation suggests that several disorders may involve a male-protective effect, raising the possibility that androgens may play a protective role in more contexts. Supporting this idea, both testosterone administration and sex reversal using an *Sry* transgene have been shown to reduce female embryonic lethality in a mouse model of MCM deficiency^92^. It is compelling to speculate that identifying a targetable downstream effector of androgen signaling could provide a potential therapeutic applicable to many NDDs.

In summary, we have identified a non-X-linked NDD where males are protected by their naturally higher androgen levels and demonstrate that this principle may be more widely applicable. These findings offer new avenues for developing targeted therapeutic interventions for PILBOS and potentially other NDDs with sex-based differences, and for exploring the role of sex-mediated penetrance in other autosomal dominant disorders.

## Materials and methods

### GeneMatcher and clinical data collection

We used the GeneMatcher platform^26^ (https://genematcher.org) to identify additional individuals carrying *CHD1* variants. Our study was specifically approved by the Icelandic Ethics Committee (Vísindasiðanefnd) and the Ethics Committee of Landspitali University Hospital. All work was completed in compliance with the Institutional Review Board of each institution and proper written consent was obtained from each patient/patient family included in the study.

### Variant Classification

Variant classification was performed by applying American College of Medical Genetics and Genomics (ACMG) guidelines using Franklin variant interpretation platform by Genoox^93^ (https://franklin.genoox.com). Missense variants were further interpreted using AlphaMissense^31^ by Google DeepMind, a model built on artificial intelligence designed to predict the effects of missense variants scoring them on a scale between 0 and 1. Missense variants were classified based on these AlphaMissense scores, with high-scoring variants flagged as deleterious (>0.761), low-scoring variants as benign (<0.331), and those in between uncertain (0.331-0.761).

### Visualization of variants

A lollipop plot was generated using Python with the Matplotlib library^94^ to visualize the distribution and frequency of variants in carried by individuals in the study.

### Phenotype scoring

A custom phenotype scoring system was designed, where those individuals carrying truncating LOF variants or putatively disruptive missense variants (categorized as deleterious or unknown by AlphaMissense) were scored based on the phenotype. One point was assigned for each of the following core phenotypic characteristics; developmental delay, speech apraxia/speech delay, hypotonia, seizures/abnormal EEG, macrocephaly, almond shaped eyes and being below 1% ile or above 99% ile in height or weight (1 point each). One point was given for intellectual disability and autism spectrum disorder, but if stated that symptoms were mild, half a point was assigned. Individuals with scarce phenotype information were excluded, i.e. less than three phenotype entries (1/20 in the truncating LOF group, 3/20 in the putatively disruptive missense group). The maximum possible score was 10 points.

### CHD1 protein production

The human *CHD1* cDNA sequence was cloned from a plasmid (Sino Biological), and the p.R618Q variant was introduced using site-directed mutagenesis (New England Biolabs). N-terminal deletions of *CHD1^WT^*and *CHD1^R^*^618^*^Q^*were generated by PCR and cloned into the pCoofy51-TwinStrep tag expression plasmid using Sequence and Ligation Independent Cloning (primer sequences provided in **Supplementary Table 9**). Protein production and purification was performed at the EMBL Protein Expression and Purification Core Facility. The proteins were produced using High Five insect cell culture, then purified using StrepTactin affinity purification and size-exclusion chromatography. The presence of CHD1 protein was confirmed by mass spectrometry.

### Nucleosome remodeling assay

The nucleosome remodeling assay was modified from the Epicypher Nucleosome Remodeling Assay protocol (https://www.epicypher.com/content/documents/protocols/epidyne-guide.pdf). Reactions were prepared by combining the previously synthesized human CHD1 proteins with N-terminal deletions (either WT or R618Q) at a concentration of 20 nM with 40 nM Epidyne Nucleosome Remodeling Assay Substrate (Epicypher, 16-4101) and 25 units of DpnII enzyme (New England Biolabs) in assay buffer (20 mM Tris pH 7.5, 50 mM KCl, 3 mM MgCl_2_, 0.1 mg/mL bovine serum albumin) to a final volume of 20 µL. Reactions were initiated by addition of 1 mM ATP, then incubated for different lengths of times at 37°C. Reactions were quenched by the addition of 20 µL assay quench buffer (10 mM Tris pH 7.5, 40 mM EDTA, 0.60% SDS, 50 µg/mL Proteinase K), then incubated at 50°C for 20 min and allowed to cool to room temperature before loading on gel. Negative controls without exposure to ATP were included for both CHD1 proteins, as well as a substrate-only control, and a positive and negative digest control (Epicypher, 23618-4101 & 23618-4100).

### Mice

The *Chd1^R^*^616^*^Q/+^* mouse line were created by Welcome Sanger (Hinxton, Cambridge) through a grant from INFRAFRONTIER 2020 to H.T.B and G.P. Mice were created by CRISPR-Cas9 to generate a mutant allele containing the missense mutation Chr17:15738534 G>A [R616Q]. In an effort to create the desired variant (R616Q), a synonymous mutation was created: Chr17:15738553 C>A (guide RNA and mutant oligo sequences in **Supplementary Table 9**). The mice carrying the synonymous variant demonstrated no obvious phenotypic difference from WT littermates, as an example we show the righting assay (**Fig. S7A**). The mice were bred and maintained according to the Institute’s standard protocols before being shipped to our facility. All animal procedures were conducted in accordance with the guidelines and regulations approved by the University of Iceland and the Icelandic Food and Veterinary Authority (license no. 2208602) and accepted by the National Expert Advisory Board on Animal Welfare. The animals were maintained in a 12 h light-dark cycle in a temperature-and humidity-controlled environment (21-23°C, ∼40% humidity), and provided with food (Altromin NIH#31 M (breeding) or Altromin 1324 (maintenance), Brogaarden) and water *ad libitum*. Cages were equipped with appropriate environmental enrichment to promote natural behaviors and well-being. Enrichment included paper nesting materials, cardboard tunnels for burrowing and exploration, and plastic shelters to provide hiding spaces.

Additionally, mice were provided with wooden chew sticks to support dental health and prevent overgrowth of teeth. Enrichment materials were replenished as needed. Mice were housed with 2-5 animals per cage. The mice were backcrossed to the C57BL/6N strain (Taconic Biosciences), then regularly crossed with WT C57BL/6N females. Genotyping was performed using tissue from ear clips with the following primers: Chd1_fwd (5’-CAGCAGGCCAGTCAACTGAGGC-3’), Chd1_rev (5’-TCTCCCAACTGCATCTGAGCACA-3’). The presence of the mutant allele was confirmed either by Sanger sequencing or by restriction digest with ClaI, which recognizes the WT but not the Chr17:15738534 G>A sequence.

### RNA extraction, cDNA synthesis and RT-qPCR

RNA was extracted by homogenization in Tri-reagent (Thermo Fisher Scientific, 15596026), then isolated using the Direct-zol RNA isolation kit (Zymo, R2062) according to the manufacturer’s instructions. cDNA was synthesized from 1 µg of RNA using the High-Capacity cDNA Reverse Transcription Kit (Thermo Fisher Scientific, 4368814) according to the manufacturer’s instructions. RT-qPCR was performed using the Luna Universal qPCR Master Mix (NEB, M3003E) according to manufacturer’s instructions on the CFX384™ Real-Time PCR Detection System (Bio-Rad). Relative quantitation was calculated according to the Pfaffl method^95^, with *Actb* used as a reference gene. To determine primer efficiencies, standard curves were prepared using 10-fold dilutions of mouse cDNA. Primer amplification factors were calculated based on 10^-1/slope^ ^of^ ^standard^ ^curve^. Each biological replicate represented an individual mouse and was performed in technical triplicate.

### Primary cell isolation and culture

Mouse embryonic fibroblasts (MEFs) were isolated from day 14.5 embryos as previously described^96^. MEFs were cultured in DMEM-F12 (Gibco, 31331093) with 10% fetal bovine serum (Gibco, A5256701), and sub-cultured when confluency reached 70-80%. Mouse cortical neuronal progenitor cells (NPCs) were isolated from day 18.5 embryos as previously described^46^. NPCs were cultured in neurobasal growth medium containing 1X B27 supplement (Thermo Fisher Scientific, 17504044), 1X Penicillin/Streptomycin (Thermo Fisher Scientific, 15140122), 1X Glutamax (Thermo Fisher Scientific, 35050038), 20 ng/mL FGF-2 (Peprotech, 100-18B), 20 ng/mL EFG (Peprotech, AF-100-15), and 2 μg/mL Heparin (MP Biomedicals, 210193125). NPCs were either sub-cultured or media was replaced every 2 days.

### Protein extraction and western blot

Cells (1 × 10^6^) were lysed in 50 µL RIPA buffer (50 mM Tris pH 8, 150 mM NaCl, 1% NP-40, 1% Sodium Deoxycholate, 0.1% SDS, 2 mM EDTA, 1 mM phenylmethylsulfonyl fluoride with 1% protease inhibitor cocktail, Sigma, P8340) on ice for 30 min, then centrifuged at 13000 rpm for 15 min at 4°C. Supernatants were mixed with 4X sample buffer (240 mM Tris pH 6.8, 8% SDS, 40% glycerol, 0.04% bromophenol blue, 5% beta-mercaptoethanol) to a 1X final concentration, then heated at 95°C for 5 min and allowed to cool to room temperature before being loaded on 8% polyacrylamide gel and electrophoresis for western blot^97^. Proteins were transferred to PVDF membranes for 2 h at 200 mA. Membranes were blocked for 1 h at room temperature in 5% bovine serum albumin (BSA, Applichem, A1391), then incubated overnight with primary antibodies (anti-CHD1, 1:1000 dilution, Cell Signaling Technologies, 4351; anti-beta-Actin, 1:1000 dilution, Abcam, ab8224). Membranes were washed 3X with tris-buffered saline with 0.1% tween-20 (TBST), then incubated 1 h with secondary antibodies (IRDye® 800CW Donkey anti-Rabbit IgG (H + L) & IRDye® 680RD Donkey anti-Mouse IgG (H + L), Licor, 926-32213 & 926-68072). Membranes were again washed 3X with TBST before visualization using an ODYSSEY® DLx (Licor). Band intensities were quantified using ImageJ Fiji software^98^.

### Mouse motor/behavioral testing

The mouse righting reflex and hindlimb strength was assessed in postnatal day 6 pups using the surface righting and hindlimb suspension tests as previously described^99^. The Morris water maze test was performed as previously described^100^. The data were analyzed using the EthoVision XT software (Noldus). For the open field test, each mouse was placed in an open field consisting of a 32 x 32 cm square enclosure open on top and allowed to roam freely while being filmed for 10 minutes. The open field enclosure was cleaned thoroughly with soap and water, then with 70% ethanol and allowed to dry completely between each test. The video data were analyzed using EthoVision XT software (Noldus), with the center of the floor area defined as an 18 x 18 cm area in the center of the open field.

### Mouse vocalization measurements

Pups at postnatal day 3-11 were temporarily removed from their dams and placed inside a soundproof box. Vocalizations were recorded over a four-minute period using the Petterson M500-384kHz USB Ultrasound Microphone with gain set to 97. Pups were then returned to their home cage. Audio recordings were processed with UltraVox software (Noldus) to count the number of vocalizations and measure the latency until first vocalization.

### Electroretinography (ERG) and Optical Coherence Tomography (OCT)

Corneal ERG responses were obtained from mice anesthetized with an IP administered mixture of ketamine (40 mg/kg) and xylazine (4 mg/kg) after 24 hours dark adaptation and then after 10 minutes light adaptation with a light-adapting background of 9 cd/m^2^. ERG recordings were performed with a Celeris D430 rodent recording system (Diagnosys LLC, Lowell, MA, USA) equipped with “bright standard” stimulators and light guide electrodes, following procedures described previously^101^. OCT scans from the eyes of anesthetized mice were obtained with a Micron IV SD-OCT system (Phoenix-Micron, Inc, Bend, OR, USA) on a rodent fundus camera. Pupils were dilated with mydriatic eye drops (phenylephrine and tropicamide) and corneas locally anesthetized with tetracaine eyedrops for both ERG and OCT examinations.

### Oxytocin ELISA

Hypothalamus tissue was manually dissected from mouse brains and lysed in RIPA buffer as above. Protein concentrations were quantified using the Pierce™ BCA Protein Assay Kit (Thermo Fisher Scientific, 23225) according to the manufacturer’s instructions. Oxytocin levels were quantified using the Oxytocin ELISA kit (Enzo, ADI-901-153A-0001). Protein lysates were normalized to a concentration of 0.4 µg/µL by dilution in the kit assay buffer and then the assay was performed according to the manufacturer’s instructions.

### MTT proliferation assay

Proliferation was assessed using the CellTiter 96® Non-Radioactive Cell Proliferation Assay MTT (Promega, G4000) according to the manufacturer’s instructions. MEFs and NPCs were seeded at a density of 1 × 10^3^ cells per well in a 96-well flat-bottomed plate. After 24 h, media was replaced with 100 µL fresh media plus 15 µL kit dye solution and plates were incubated at 37°C (MEFs for 3 h, NPCs for 4 h). The reaction was stopped by the addition of 100 µL stop solution and incubated for 1 h at room temperature protected from light. Absorbance was read at 570 nm and results were normalized by subtracting the background signal.

### Cell cycle analysis

Cells were harvested and pelleted, then pellet was resuspended in 400 µL ice-cold PBS. While gently vortexing, 800 µL ice-cold 100% ethanol was added to a final concentration of 66% for fixation. After washing in PBS, cells were stained with 0.2 µg/mL DAPI (AAT Bioquest, ABD-17511) diluted in PBS, with 100 µg/mL RNase A (Abcam, ab139418) and incubated at 37°C for 30 min protected from light. Staining was measured by flow cytometry using the FACS Calibur (BD, for MEFs) or Attune NxT (Thermo Fisher Scientific, for NPCs). Data were analyzed with FlowJo software (v.10.8.1) with gating to include only live single cells (**Fig. S7B**).

### EdU DNA synthesis assay

DNA synthesis was measured using the Click-iT™ Plus EdU Alexa Fluor™ 647 Flow Cytometry Assay Kit (Thermo Fisher Scientific, C10634) according to the manufacturer’s instructions. Cells were incubated with 10 µM EdU for 2h before harvesting, fixation and Click-iT reaction. EdU incorporation was assessed by flow cytometry using Attune NxT (Thermo Fisher Scientific).

### Annexin V apoptosis assay

Apoptosis was quantified using the FITC Annexin V Apoptosis Detection Kit I (BD, 556547) according to the manufacturer’s instructions. Staining was measured by flow cytometry using the Attune NxT (Thermo Fisher Scientific) and FlowJo software (v.10.8.1) was used for analysis.

### RNA sequencing

NPCs obtained from 4 individual embryos per genotype from 2 separate litters were seeded (passage 3) 1.5 × 10^5^ cells per well in a 6-well plate coated with poly-D-lysine and laminin. After 3 days of culture, cells were treated with DMSO vehicle control or 10 nM dihydrotestosterone (Selleckchem, S4757) for 6 h, then RNA was extracted as described above. RNA quality and integrity was assessed using a Nanodrop spectrophotometer and a Bioanalyzer instrument with the RNA 6000 Nano Kit (Agilent, 5067-1511). Only samples with RNA integrity (RIN) scores above 8 were used for library preparation. PolyA-enriched RNAseq library construction and Illumina paired-end sequencing was performed by Novogene, to obtain 30 million paired-end 150 bp reads for each sample. Reads were pseudoaligned to the GRCm39 transcriptome using Kallisto using paired-end mode and 100 bootstraps, then prepared for differential expression analysis using the Tximport package in R, using the TxDb.Mmusculus.UCSC.mm10.ensGene package to assign reads to genes. Differential expression analysis was performed using DESeq2, keeping only transcripts with at least 10 counts. For the DMSO vs DHT treatment comparison, we used a design matrix that accounted for embryo as well as genotype and treatment, since both treatments were each applied to the same embryo. Since the p-value histograms displayed a right-skewed distribution, which can indicate non-random noise or inappropriate assumptions, we utilized the DESeq2 z-score data as input for the FDRtool package to achieve more accurate false discovery rate estimations, and used these corrected p-values and q-values for further analysis. Overlaps were calculated by Fisher’s exact test using the GeneOverlap package in R. Overrepresentation analysis was performed using WebGestalt^102^ with the functional database settings, “GeneOntology: CellularComponent” and “Pathway: KEGG”.

### Mutational signature analysis

The method used for variant discovery in the RNAseq data was adapted from the GATK^103^ Best Practice Workflow for RNAseq short variant discovery (https://gatk.broadinstitute.org/hc/en-us/articles/360035531192-RNAseq-short-variant-discovery-SNPs-Indels). Raw RNAseq data were aligned to the GRCm39 v. 112 mouse genome using STAR^104^ with two-pass mode, with bam output files sorted by coordinate. Duplicates were marked in bam files using Picard (https://broadinstitute.github.io/picard/) then with GATK tools^103^, SplitNCigarReads was used to re-format intron-spanning alignments, BaseRecalibrator, ApplyBQSR, and AnalyzeCovariates were used for base quality recalibration. For variant discovery, first variants were called on the WT samples using GATK Mutect2 in tumor-only mode to obtain variants that were then used to assemble a panel of normals using the CreateSomaticPanelOfNormals command. Variants were then called in each sample against the panel of normals using Mutect2. Variants common to at least 12 samples out of 16 were identified using the BCFtools^105^ -iSec command, then filtered out of the VCF files. VCF files were read into R using the MutationalPatterns package^106^ and limited to variants found within exons using the TxDb.Mmusculus.UCSC.mm39.refGene package. Indels were assessed using the get_indel_context() command. Mutational signatures were fit to known COSMIC signatures^47^ using the fit_to_signatures() command.

### Orchiectomy and testosterone implant surgeries

Surgeries were performed on mice at postnatal day 15. Mice were anesthetized with 40 mg/kg ketamine and 4 mg/kg xylazine intra-peritoneally, as well as 7 mg/kg lidocaine subcutaneously at the incision site.

In male mice, two abdominal incisions were made, one for each testis. Testicular tissue was pulled through the incision by the epididymal fat to expose the testis, and then the testicular blood vessels were crushed using hemostats with heat applied to prevent hemorrhage. The testes were then excised, and the incision closed with absorbable sutures (Vicryl, 4-0, Ethicol, V310H). For post-operative analgesia, male mice were treated with 0.1 mg/kg buprenorphine. In female mice, a small incision was made in the back of the neck and the testosterone 60-day slow-release silastic implant (Belma Technologies, T-M/60, Batch No. M60T1711-03NB) was placed under the skin. The incision was closed with an absorbable suture.

### Testosterone treatment of dams

Testosterone propionate (T, MedChemExpress, HY-B1269) was dissolved in corn oil to a stock concentration of 5 mg/mL by heating for 1 h at 50°C with frequent vortexing. Pregnant dams were injected with 20 mg/kg T or 4 µL corn oil per gram body weight subcutaneously in the hindleg once per day from embryonic day 8.5-17.5. On embryonic day 18.5, dams were euthanized by cervical dislocation and embryo telencephalon tissue was used to isolate nuclei. Embryos were obtained from 3-4 litters for each treatment.

### Nuclear isolation for flow cytometry

The telencephalon was dissected from embryo brains and placed in ice-cold homogenization buffer (5 mM calcium chloride, 3 mM magnesium acetate, 10 mM Tris pH 7.8, 0.17% beta-mercaptoethanol, 320 mM sucrose, 1 mM EDTA, 0.1% IGEPAL CA-630) and homogenized using 2 mL Dounce homogenizer.

Homogenate was transferred to 1.5 mL centrifuge tube and centrifuged 8000 rpm for 30 seconds to pellet nuclei. Nuclei were resuspended in 200 µL FACS buffer and stained with anti-Ki67 (1:500, Abcam, ab16667) for 20 min at room temperature. Samples were stained with Alexa fluor-647 anti-rabbit secondary (1:1000, Thermo Fisher Scientific, A-21244) and 0.2 µg/mL DAPI (AAT Bioquest, ABD-17511) for 30 min at room temperature protected from light. Nuclei were passed through a pre-wet 100 µm filter, then fluorescence was measured by FACS using the SH800S flow cytometer (Sony). Data were analyzed using FlowJo software (v.10.8.1).

### Human population variant analysis

The list of all genes with available pLI scores was downloaded from the gnomAD v4.1 website^30^ (gnomad.v4.1.constraint_metrics.tsv). This list was filtered to keep only genes with pLI score > 0.9. The gnomAD v4.1 exome hail table data (“gnomad.exomes.v4.1.sites.ht”) was downloaded from the gnomAD website^30^. Variants assigned to genes of interest were then selected, and the data were filtered to exclude sex chromosomes. Then the variant filters “AC0” (variants whose allele count is zero after filtering out low-confidence genotypes) and “AS_VSQR” (variants that failed allele-specific Variant Quality Score Recalibration filtering thresholds of -4.0598 for SNPs and 0.1078 for indels) were applied. We then limited to missense variants that occurred at < 1% adjusted allele frequency at sites with at least 100 000 allele number (AN) counts.

Analysis of data from the UK Biobank resource was performed at deCODE Genetics using the UK Biobank resource, application number 56270. The filtered list of genes with pLI score > 0.9 previously obtained from gnomAD was used to extract rare (MAF < 1%) missense variants from the UK Biobank WGS dataset.

For data extracted from all datasets, we calculated the aggregated alternate allele frequency per gene as the sum of the alternate allele counts (AC) divided by the sum of the allele number (AN) counts for each sex. We then used the two-proportions z-test using the prop.test() function in R to compare the aggregated alternate allele frequencies for each sex. The *p*-values were adjusted using the Benjamini-Hochberg correction. The frequency difference (FrqDiff) was calculated by subtracting the allele frequency for females from that of males (male – female). Overrepresentation analysis was performed using WebGestalt^102^ with the functional database settings “Gene ontology: Molecular Function” and “Pathway: Reactome”. Overlaps were calculated by Fisher’s exact test using the GeneOverlap package in R.

Analysis in the All of Us Research Program’s Controlled Tier Dataset version 8, were based on the exome sequencing data in a maximal set of unrelated samples^54^. We identified biallelic missense variants in *CHD1* from the columns “consequence” and “gene_symbol” in the variant annotation table provided in the All of Us data release. Allele frequencies were computed for these variants in the exome sequencing data and variants with MAF ≥ 1% were subsequently filtered out. After identifying individuals carrying at least one rare missense variant in *CHD1*, we performed logistic regression in R where sex at birth (male or female) was the dependent variable and *CHD1* rare missense variant carrier status (carrier or non-carrier) was the explanatory variable (n = 380,281, male-fraction = 39.8%, carrier-fraction = 2.3%).

## List of Supplementary Tables

Supplementary Table 1. Phenotypes observed in the PILBOS cohort with missense variants.

Supplementary Table 2. Phenotypes observed in the PILBOS cohort with LOF and potential splice-site variants.

Supplementary Table 3. Mendelian ratios of *Chd1^R^*^616^*^Q/+^* mice and WT littermates.

Supplementary Table 4. Differentially expressed genes in NPCs from *Chd1^R^*^616^*^Q/+^* and wildtype mouse embryos.

Supplementary Table 5. Differentially expressed genes in NPCs from *Chd1^R^*^616^*^Q/+^* mouse embryos, treated with DMSO vehicle control or dihydrotestosterone.

Supplementary Table 6. A list of genes from gnomAD non-UKB dataset - all autosomal genes with pLI > 0.9 with test for male or female overrepresentation in rare missense variants

Supplementary Table 7. A list of genes from UK Biobank WGS dataset - all autosomal genes with pLI > 0.9 with test for male or female overrepresentation in rare missense variants

Supplementary Table 8. A list of genes that show sex-specific skewing of missense variants in gnomAD and UK Biobank.

Supplementary Table 9. Guide RNA and oligo sequences used to create the mouse model, primer sequences from the current study.

## Data availability

Raw sequencing data have been deposited in the Gene Expression Omnibus (GEO) under accession number GSE281601. Data will be made publicly available upon peer-reviewed publication of the manuscript.

This study used data from the All of Us Research Program’s Controlled Tier Dataset version 8, available to authorized users on the Researcher Workbench.

## Supplementary figures

**Supplementary Figure 1:**
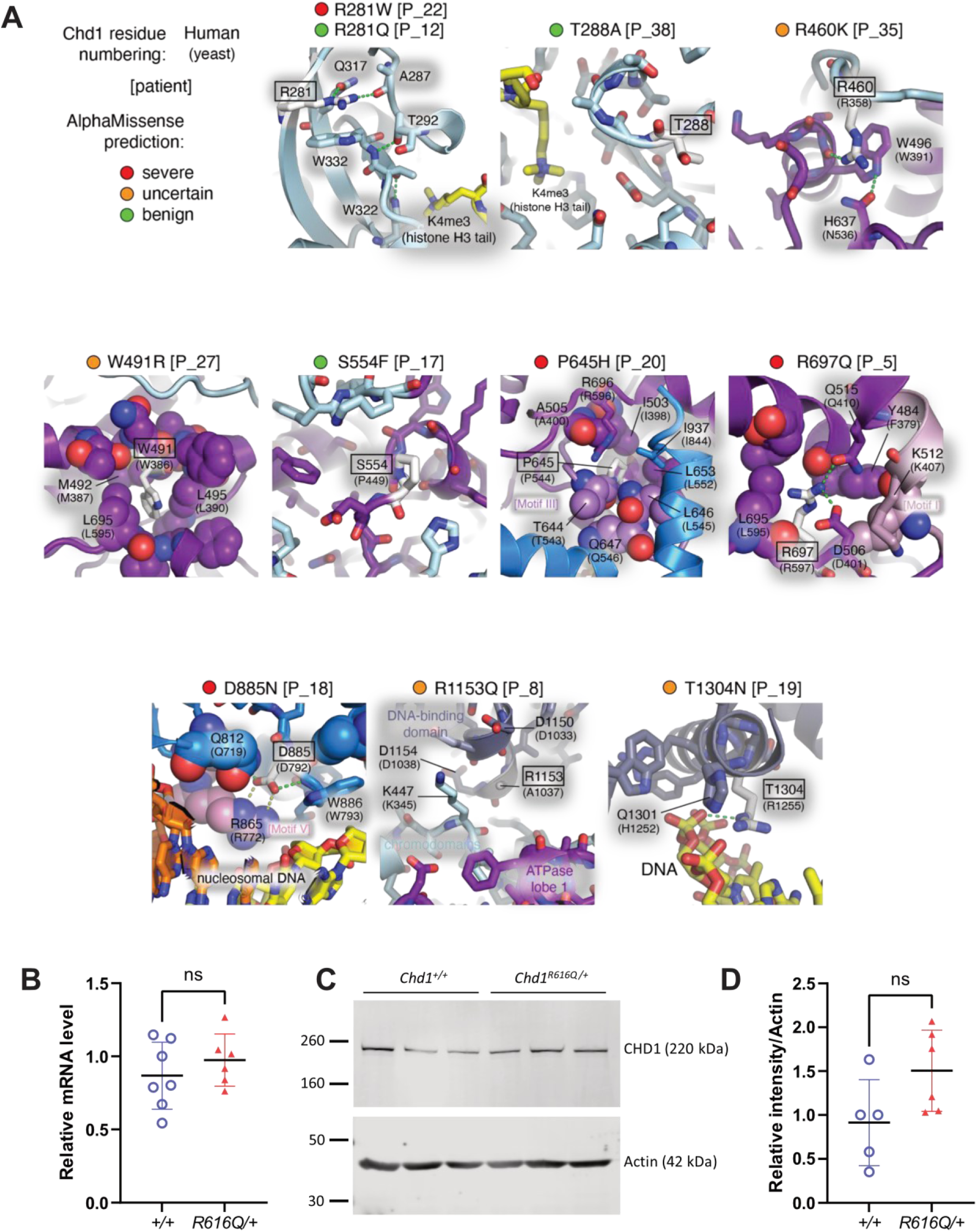
The effects of CHD1 variants. (A) Variant positions (white, boxed) were analyzed using Chd1 crystal and cryoEM structures from yeast (PDB code 7TN2, 3TED; with yeast amino acid positions shown in parentheses beneath corresponding human positions) and human (2B2W). In each case, neighboring residues that may be affected by the missense change are highlighted as well. Those considered to be deleterious are often part of, contacting, or near conserved helicase motifs. (B RT-qPCR results showing *Chd1* mRNA levels in tissue obtained from the adult cortex of *Chd1^R^*^616^*^Q/+^* mice and WT littermates. (C) A representative western blot showing CHD1 protein levels in mouse embryonic fibroblasts (MEFs), quantified in (D). ns: not significant.

**Supplementary Figure 2:**
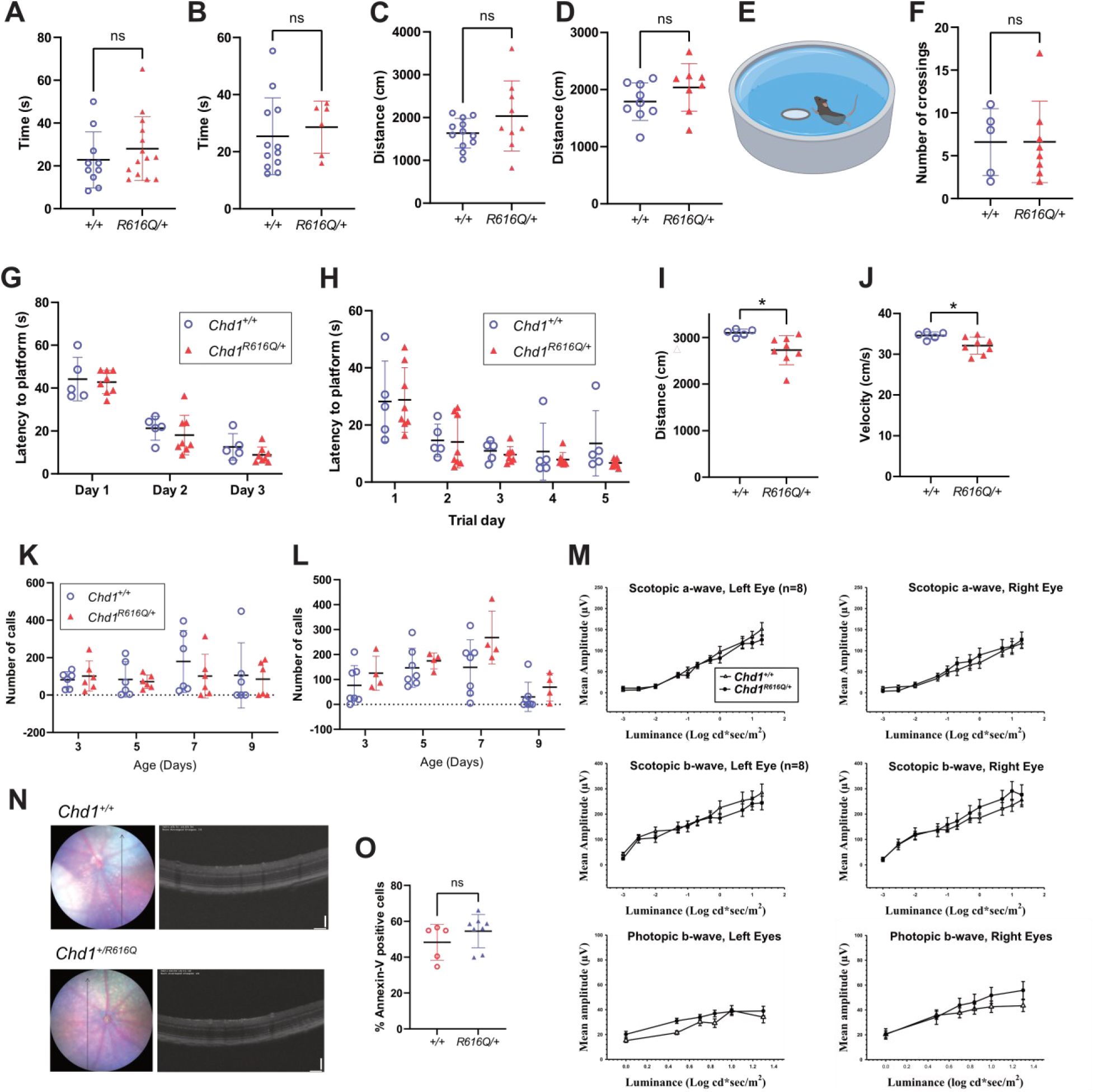
Additional phenotyping of the *Chd1^R^*^616^*^Q/+^* mice. Time spent in suspension during the hindlimb suspension test in (A) females (WT: *n* = 10, *R616Q/+*: *n* = 14) and (B) males (WT: *n* = 12, *R616Q/+*: *n* = 6). (C) Distance travelled in an open field during the 10-minute period for (C) females (WT: *n* = 12, *R616Q/+*: *n* = 9) and (D) males (WT: *n* = 9, *R616Q/+*: *n* = 8). (E) A diagram illustrating the basis of MWM swimming task, females only (WT: *n* = 5, *R616Q/+*: *n* = 8). (F) Quantification of the number of platform crossings of female *Chd1^R^*^616^*^Q/+^* mice and WT littermates during the hidden platform test on the final day of the MWM task, used as a measure of visuospatial learning and memory. (G) Time latency to visible platform. (H) Time latency to hidden platform. Quantification of (I) distance travelled and (J) swimming velocity of female *Chd1^R^*^616^*^Q/+^* mice and WT littermates during the hidden platform test of the MWM. Quantification of the total number of pup calls over a 4-minute period upon separation from their dams at postnatal day 3, 5, 7 and 9 in (K) females (WT: *n* = 6, *R616Q/+*: *n* = 6) and (L) males (WT: *n* = 7, *R616Q/+*: *n* = 4). (M) Mean amplitudes of dark-adapted ERG a- and b-waves and light-adapted b-waves of *Chd1^R^*^616^*^Q/+^* mice and WT littermates. (N) Sample OCT scans from the eyes of a *Chd1^R^*^616^*^Q/+^* mouse and a WT littermate. Locations of the scans on the fundi are indicated by the black vertical arrows in the adjacent fundus images. (O) The percentage of annexin V positive NPCs in culture as measured by flow cytometry (WT: *n* = 5, *R616Q/+*: *n* = 8). *p < 0.05, ns = not significant.

**Supplementary Figure 3:**
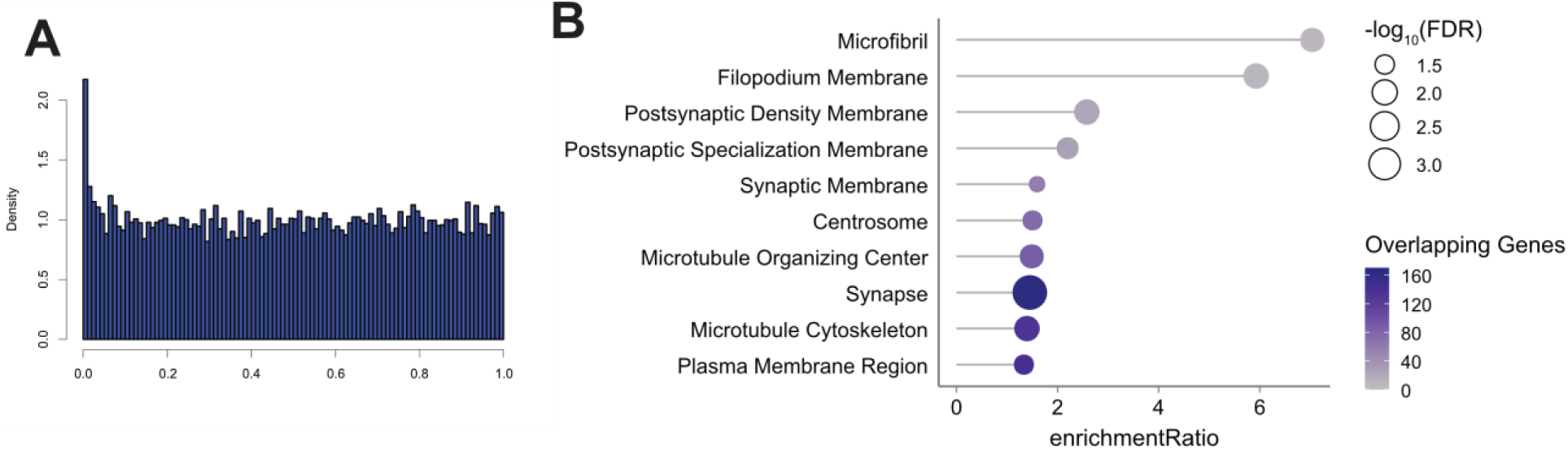
Quality measures and enriched pathways from RNA-Seq. (A) Histogram of p-values for the comparison *Chd1^+/+^* vs *Chd1^R^*^616^*^Q/+^* bulk RNAseq in NPCs (*n* = 4 per genotype). (B) Cellular component gene ontology overrepresentation analysis using genes with the top 10% most significant *p*-values for the comparison of *Chd1^+/+^*vs *Chd1^R^*^616^*^Q/+^*.

**Supplementary Figure 4:**
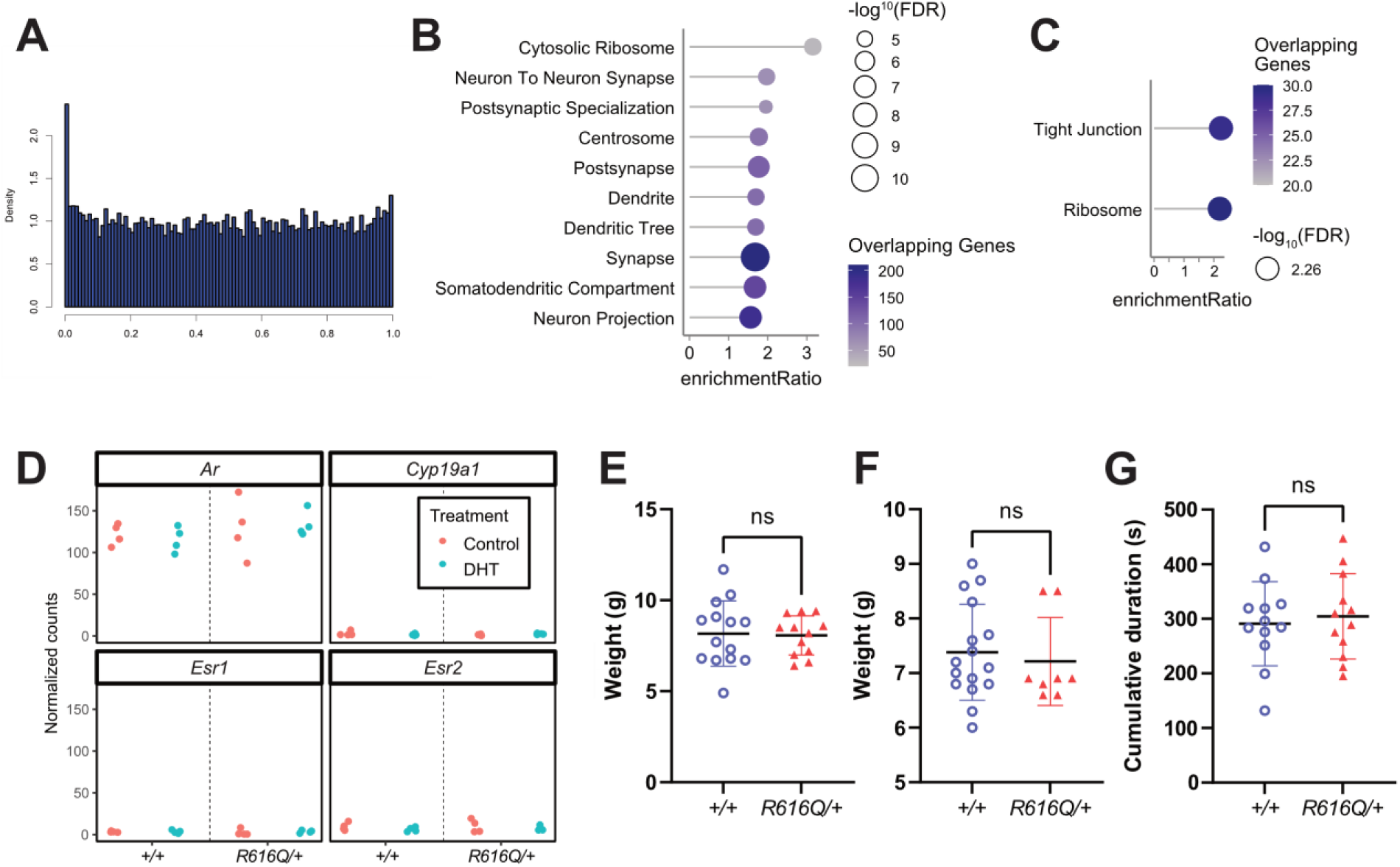
Additional data regarding the effect of DHT on gene expression in the two genotypes. (A) A histogram of *p*-values for the comparison of DMSO control vs DHT treatment only in *Chd1^R^*^616^*^Q/+^*NPCs (*n* = 4 per treatment). (B) Cellular component gene ontology overrepresentation analysis using genes with the top 10% most significant *p*-values for the comparison of DMSO control vs DHT treatment only in *Chd1^R^*^616^*^Q/+^*NPCs. (C) KEGG pathway gene ontology overrepresentation analysis for the comparison of DMSO control vs DHT treatment only in *Chd1^R^*^616^*^Q/+^* NPCs. (D) Normalized count data for the genes *Ar*, *Cyp19a1*, *Esr1* and *Esr2*. (E) Weights of male mice at P15 prior to orchiectomy. (WT: *n* = 14, *R616Q/+*: *n* = 12). (F) Weights of female mice at P15 prior to testosterone implantation. (WT: *n* = 16, *R616Q/+*: *n* = 8). (G) Cumulative duration of time spent in the center of the open field for orchiectomized male mice at 3 months of age (*n* = 12 per genotype). ns = not significant.

**Supplementary Figure 5:**
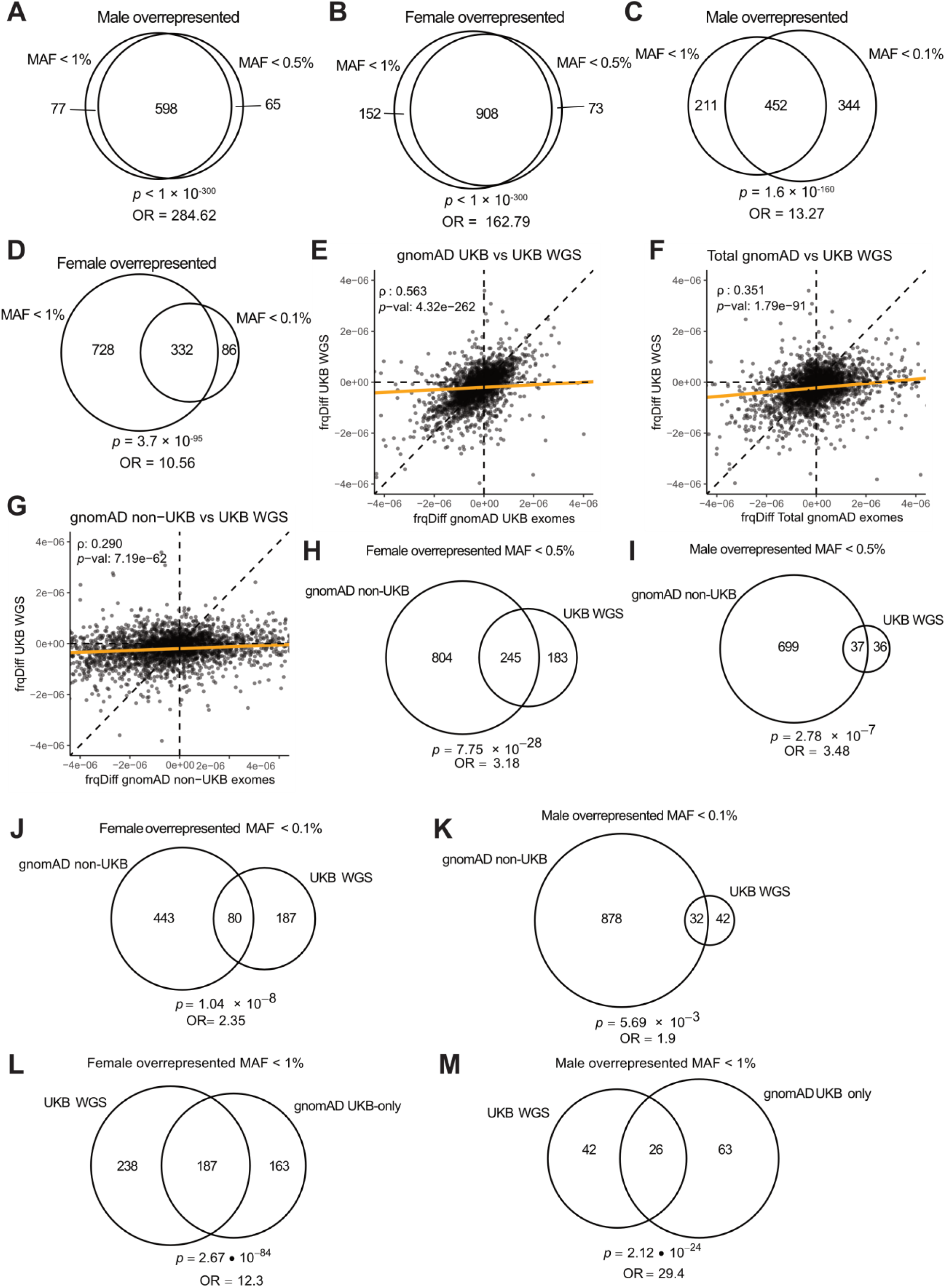
Quality control for sex differences in rare missense variants in gnomAD and UK Biobank. Overlapping genes with significant sex overrepresentation of missense variants in gnomAD exome data identified using MAF cutoffs of < 0.5% and <1 in (A) males and (B) females. Overlapping genes with significant sex overrepresentation of missense variants identified using MAF cutoffs of < 1% and <0.1% in (C) males and (D) females. Male – female aggregated allele frequency differences (frqDiff) for rare (MAF < 1%) missense variants in (E) gnomAD UK biobank exome sequencing samples compared to UK Biobank WGS samples, (F) total gnomAD exomes dataset compared to UK Biobank WGS samples, and (G) gnomAD non-UK Biobank exomes samples compared to UK Biobank WGS samples. Spearman’s correlation (ρ) and *p*-value shown for each plot. Each dot represents one gene. Overlapping genes with sex overrepresentation for missense variants with MAF < 0.5% in gnomAD non-UKB and UKB WGS datasets in (H) females and (I) males. Overlapping genes with sex overrepresentation for missense variants with MAF < 0.1% in gnomAD non-UKB and UKB WGS datasets in (J) females and (K) males. The overlap of sex-biased genes identified from variants with MAF < 1% and *p*-value < 0.05 in the gnomAD UKB-only dataset compared to the UKB WGS dataset for genes with (L) male overrepresentation and (M) female overrepresentation. All *p-*values and odds ratios calculated using Fisher’s exact test, with a background of 3140 genes (the number of autosomal genes with pLI > 0.9).

**Supplementary Figure 6:**
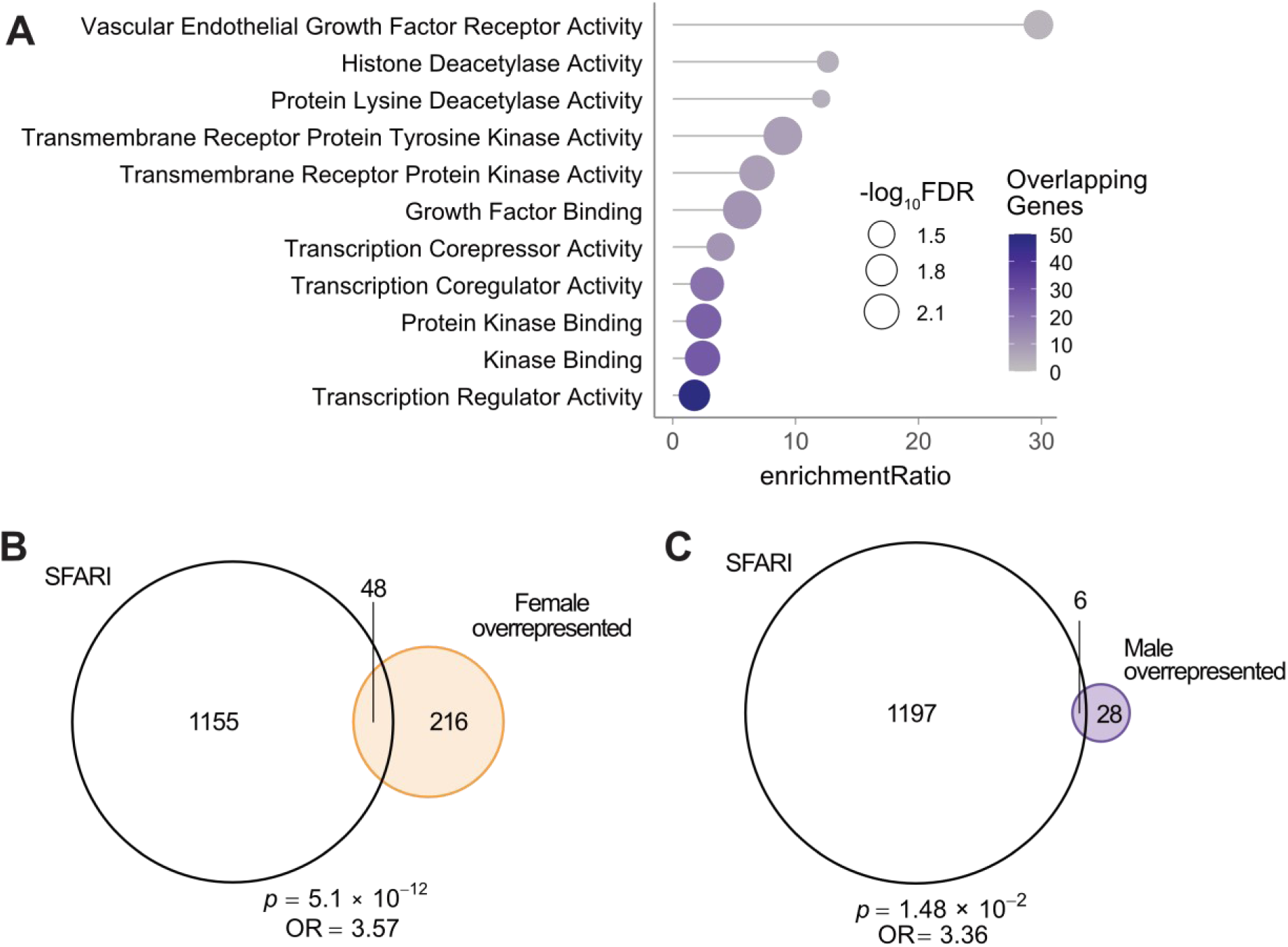
Functional analysis of genes that show a sex bias in our study. (A) Molecular function gene ontology overrepresentation analysis of genes with female overrepresentation in gnomAD and UKB. Venn diagrams depicting the overlap of SFARI genes with our identified lists of genes with (B) female or (C) male overrepresentation in gnomAD and UKB. The *p* values and odds ratios for the overlaps were calculated using Fisher’s exact test using a background of 19980 genes (number of protein coding genes in GENCODE hg38).

**Supplementary Figure 7:**
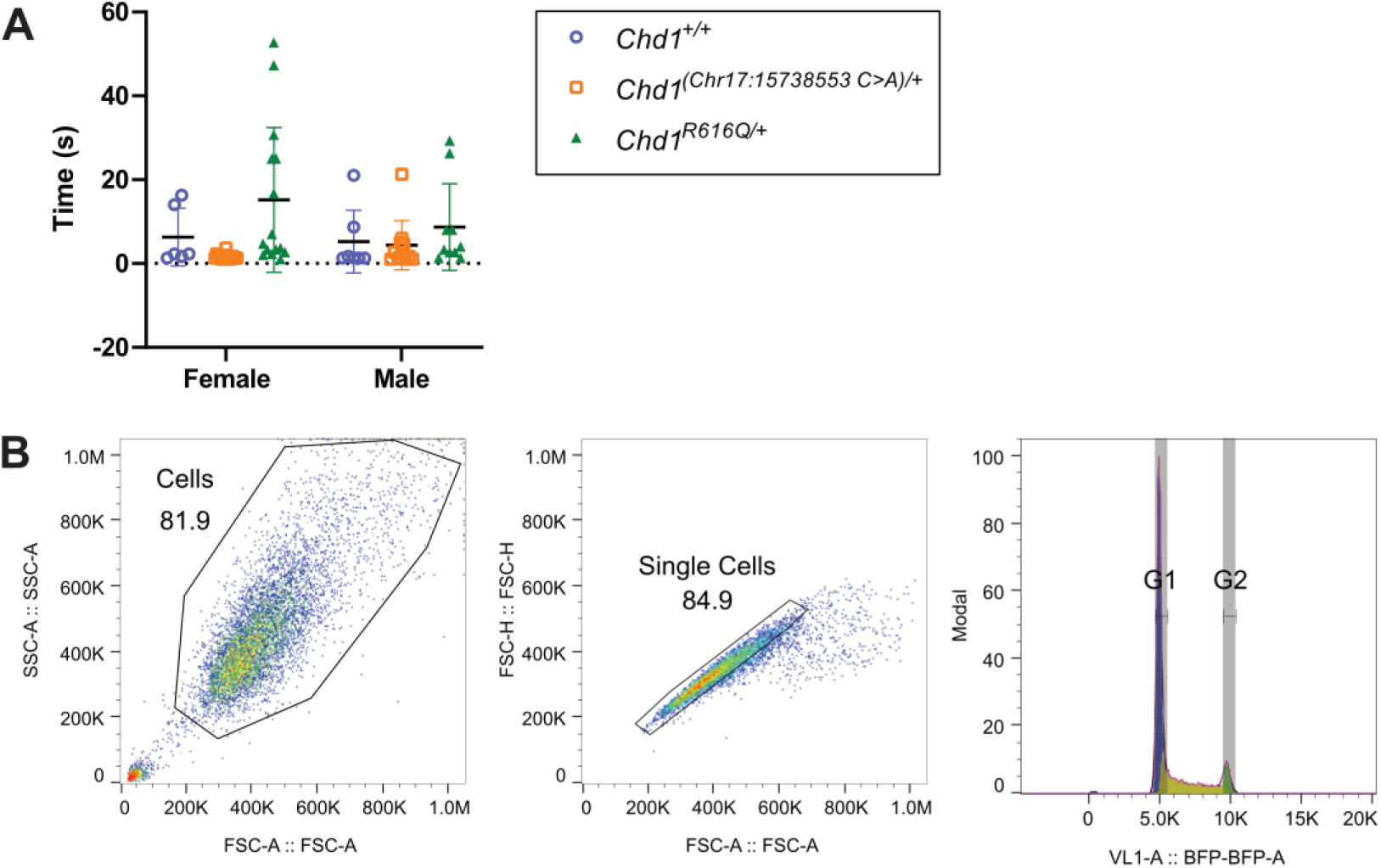
*Chd1* synonymous mutation and flow cytometry gating for single cells using NPCs for cell cycle analysis. (A) Quantification of righting reflex times in female (WT: *n* = 6, *Chr17:15738553 C>A/+*: *n* = 14, *R616Q/+*: *n* = 15) and male (WT: *n* = 7, *Chr17:15738553 C>A/+*: *n* = 11, *R616Q/+*: *n* = 10) *Chd1^R^*^616^*^Q/+^*, *Chd1^(Chr^*^17^:^15738553^ *^C>A)/+^*, and WT littermates at postnatal day 6. The plot includes data depicted in **Fig. 2C**). (B) Flow cytometry gating for single cells and cell cycle analysis using DAPI staining of NPCs.

## Supplementary Tables

**Supplementary Table 3:** Altered Mendelian ratios of *Chd1^R^*^616^*^Q^* mice

| Genotype | Expected | Observed | Females | Males |
| --- | --- | --- | --- | --- |
| +/+ | 39 | 39 | 21 | 18 |
| R616Q/+ | 78 | 57 | 30 | 27 |
| R616Q/R616Q | 39 | 0 | 0 | 0 |

**Supplementary Table 9:**
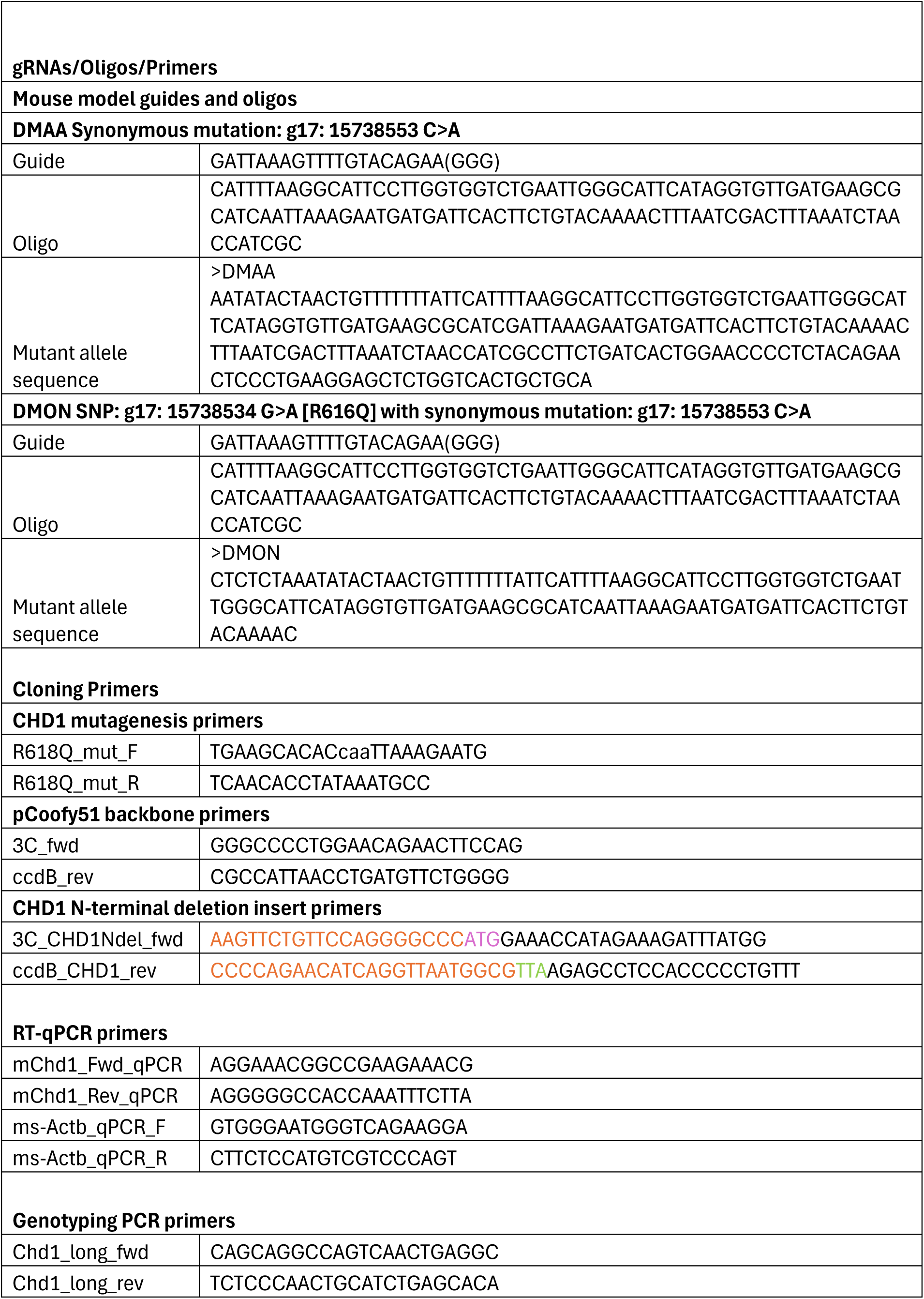
Guide RNAs, oligos and primer sequences from the study.

## Acknowledgements

We would like to thank all the individuals with PILBOS diagnosis and their families that participated in this study. We would like to thank the European mouse project. We thank the Wellcome Trust Sanger Institute Mouse Genetics Project (Sanger MGP) and its funders for providing the mutant mouse line *Chd1^R^*^616^*^Q/+^* (15738534 G>A [R616Q] with synonymous mutation: 15738553 C>A). Funding information may be found at www.sanger.ac.uk/mouseportal and associated primary phenotypic information at www.mousephenotype.org. This research has been conducted using the UK Biobank Resource under Application Number 56270. We gratefully acknowledge All of Us participants for their contributions. We also thank the National Institutes of Health’s All of Us Research Program for making available the participant data examined in this study. We thank the National Center for Medical Genomics, Czech Republic (LM2023067) for WES analyses.

## Funding

H.T.B. is funded by the Louma G. Foundation. This project was also specifically funded by a 3-year grant from Landspitali Science Fund (H.T.B.). The mouse model (*Chd1^R^*^616^*^Q/+^*) creation was funded by a grant from INFRAFRONTIER 2020. The behavioral core equipment used for behavioral testing was funded by a grant from the Icelandic Infrastructure Fund (191708-003). E.E.E. is funded by the Simons Foundation (RFA 810018EE). L.N., D.G. and S.K. were supported by grant NU23-07-00281 from the Ministry of Health of the Czech Republic. L.N. and S.K. were supported from the project MULTIOMICS_CZ (Programme Johannes Amos Comenius, Ministry of Education, Youth and Sports of the Czech Republic/ID Project CZ.02.01.01/00/23_020/0008540) – Co-funded by the European Union. This work was also supported, in part, by the National Natural Science Foundation of China (82201314 and 82471194) and the “Fundamental Research Funds for the Central Universities” starting fund (BMU2022RCZX038) to T.W. G.D.B. is funded by NIH grant R01-GM084192.

## Author contributions

H.T.B., G.P. and K.J.A. conceived the study. H.T.B. acquired funding for the study. K.J.A., S.T.H., and T.E. performed all the experimental work. E.T.T. and H.T.B. performed clinical collection and characterization of clinical variants. G.D.B. and I.M.N. did computational modelling of missense variants. K.J.A., S.B., A.H., and K.S. performed human population variant analysis. M.A., N.A., B.J.B., M.B., J.F.B., S.B., B.C., A.S.A.C, C.D.B.B., E.E., E.C.E., J.A.F., L.F., M.F., N.F., C.W.G., G.G., D.G., M.G., J.R.H., K.H., T.H., S.H., K.J., J.A.J., S.K., C.K., M.A.K., P.L., T.L., M.M., A.M., E.N., M.N., L.N., K.P., C.P., R.P., P.P., N.R., N.H.R., C.R., J.Roohi., J.Rosenfeld., M.S., C.S., Z.S., I.T., S.T., D.V., C.V., J.V., A.V., T.W., K.W., I.M.W, and M.G. provided clinical data.

## Competing interests

Dr. Bjornsson is Founder of KALDUR therapeutics and a consultant for Mahzi therapeutics. S.B. is a part of a working group called Alzheimer diagnostics that has received a grant from the Icelandic Technology Development Fund. I.M.W. is an employee of and may own stock in GeneDx. E.E.E. is a scientific advisory board (SAB) member of Variant Bio, Inc. K.S. and A.H. are employees of deCODE genetics, a subsidiary of Amgen. The Department of Medical and Molecular Genetics at Baylor College of Medicine receives revenue from clinical genetic testing completed at Baylor Genetics Laboratories.

## Declaration of generative AI and AI-assisted technologies in the writing process

During the preparation of this work the authors used Chat GPT-4 in order to improve language and readability, and for troubleshooting and refining R and python scripts. After using this tool/service, the authors reviewed and edited the content as needed and take full responsibility for the content of the publication.

## Notes

### Author Declarations

The Icelandic Ethics Committee (Visindasidanefnd) and the Ethics Committee of Landspitali University Hospital gave ethical approval for this work.

